# Dynamics of early gut microbiota maturation in extremely preterm infants in relation with neurodevelopment at 2 years

**DOI:** 10.1101/2025.03.17.25324095

**Authors:** Thomas Abrahamsson, Erik Wejryd, Eva Sverremark-Ekström, Maria C Jenmalm, Magalí Martí

## Abstract

Preterm birth yields survivors with heightened susceptibility to long-term neurological deficits, with severity being linked to gestational age. The mechanisms behind neurodevelopmental impairment remain elusive, although research underscores the role of the gut microbiome in regulating the gut-brain axis. In this prospective observational cohort study (PROPEL-study follow-up) we evaluated whether modulation of the gut microbiota by *Limosilactobacillus reuteri* supplementation to 105 extremely preterm infants with extremely low birth weight (EPT-ELBW), as well as their microbiota development during the first month of life, are associated with neurodevelopment at two years of age. The gut microbiota was characterized with 16S amplicon sequencing, and neurodevelopment was assessed with clinical examination and the Bayley-III score. Our results indicate that specific microbiota composition constellations are associated with impaired neurodevelopment, as indicated by alpha- and beta-diversity being discriminative for neurodevelopment, but not a single bacterium, with increased bacterial diversity being associated to normal neurodevelopment. Microbial maturation over the first month of life was also discriminative for neurodevelopment, where higher abundance of *E. coli* and *Enterococcus* in relation to *Enterobacter* was associated to impaired neurodevelopment. In conclusion, the dynamics of gut microbiota maturation during early of life may have an impact on neurodevelopment in EPT-ELBW infants.

## INTRODUCTION

Preterm birth is a leading cause of infant mortality and often results in morbidity and disability among survivors (1). While mortality rates have decreased during the last decade (2), the survivors often develop long-term neurological deficits with cognitive and behavioral problems (3,4). The severity of the impairment is inversely associated with gestation age, where extremely preterm (EPT; born before 28 completed gestational weeks (gw)) infants are at higher risk of suffering from neurodevelopment disabilities (5,6). The long-term neurological disabilities in adolescence and adulthood are mainly cognitive impairment, learning and language disorders, attention deficit hyperactivity disorder (ADHD), intellectual disability and autism (4,5,7).

Neurodevelopmental impairment after EPT birth is common. The French Epipage-2 trial found that 50% of infants born in gw 24 – 26 exhibited a moderate or severe neurodevelopmental impairment (< 2 SD below reference). The communication domain was most severely affected (8). In British three-year-old children born in gw 22 – 26 during 2006 a moderate or severe impairment was found among 25% of the survivors (9). Swedish infants born before gw 27 during 2004 – 2007 assessed at 2.5 years corrected age had moderate to severe impairment in 11% (cognition) and 16% (language) of the cases (6). A Japanese study of infants born 2003 – 2014 showed that the level of neurodevelopmental impairment among EPT infants was not decreasing over time (10).

The mechanisms leading to neurodevelopment impairment remain unclear, although during the last decade, it has become apparent that the gut microbiome is a key player regulating the gut-brain axis function (11). During the postnatal period, early stages of neurodevelopment occur in parallel with the gut microbiota development (12,13), and increasing evidence indicates that the early life microbiota contributes to the bidirectional gut-brain axis communication and consequent brain development and maturation regulation (12,14–16). It is during these critical early stages of development, when the gut microbiota composition as well as its maturation in preterm infants differs from the term-infants (12,17–21). Nonetheless, EPT infants remain poorly investigated, although a recent study showed that an aberrant development of the gut-microbiota-immune-brain axis during the first months of life, may be associated with neurophysiological development resulting in brain injury during the neonatal period (22).

In contrast to EPT, more studies have assessed the effects of probiotic supplementation (mainly *Bifidobacterium sp, Lactobacillus sp,* and/or *Limosilactobacillus sp* to preterm infants in relation to neurodevelopment and, generally, no effects of probiotic supplementation in relation to neurodevelopment have been observed (23,24). However, Romeo et *al*. (25) showed that preterm infants supplemented with *L. reuteri* or *Lacticaseibacillus rhamnosus* had lower incidence of abnormal neurological outcomes at 1 year of age, and Jacobs et *al*. (26) found preterm infants supplemented with *B. infantis, B. lactis,* and *Streptococcus thermophilus* to have lower incidence of sensorial hearing loss at two years corrected age.

Altogether, the results of previous studies suggest that the infant demographics (gestational age at birth, birth weight, study site), the age when gut microbiota was analyzed, time/duration/strain(s) of probiotic supplementation, as well as infant age for neurodevelopment assessment impact the results. Therefore, it is important to have a good coverage of all different infant cohorts, and thus more studies including extremely preterm infants. In a previous clinical trial (PROPEL), we have showed that daily supplementation during the neonatal period with *L. reuteri* DSM 17938 to extremely preterm infants with extremely low birth (EPT-ELBW), modulates their gut microbiota (27), and is associated with improved head growth during the first month of life (28) as well as better language development at two years corrected age (29). Therefore, in this study we sought to evaluate (a) whether modulation of the gut microbiota by *L. reuteri* supplementation and (b) microbiota development during the first month of life, are associated with better neurodevelopment at 24 months corrected age, including language development, cognition development, motor-skills development, and overall neurodevelopment impairment (NDI).

## MATERIALS AND METHODS

### Cohort

This study was part of a prospective randomized, double-blind, placebo-controlled, multi-center trial evaluating the effect of daily supplementation with the probiotic *L. reuteri* DSM 17938 in EPT-ELBW. The trial was designed to evaluate whether daily *L. reuteri* DSM 17938 supplementation improved enteral feeding tolerance. In brief, 134 EPT-ELBW infants (born between gestational week 23+0 and 27+6, weighing ≤ 1000 g) were enrolled and daily supplemented with either *L. reuteri* DSM 17938 (1.25 x 10^8^ bacteria (0.2 mL drops) or placebo from birth to post-menstrual week 36. The infants were fed exclusively with breast milk (mother’s own milk and/or donor milk) until they had reached a weight of at least 2,000 g. For a detailed description of the trial and the clinical outcomes see Wejryd et al. (28).

### Two years follow-up

EPT infants undergo a routine follow-up in Sweden according to national guidelines. This includes an assessment with Bayley Scales of Infant and Toddler Development, 3^rd^ edition (Bayley-III) (30). From the PROPEL-trial, 110 infants participated in the follow up and were assessed by psychologists in Linköping and Stockholm. The composite index score for cognition, motor and language scales were used. A neonatologist or pediatrician collected information on medical history, physical examination, auxology, standardized neurologic examination, assessment of general appearance of gross- and fine motor function and whether the child was perceived to have normal general development. Data was reported to the Swedish Neonatal Quality Register (www.snq.se) and retrieved from the register for the study subjects. The trial was reviewed and approved by the Ethics Committee for Human Research in Linköping (Dnr 2012/28-31, 2016/503-32, 2019-04975). A composite of neurodevelopment impairment was also calculated on data from the follow-up, i.e., Bayley-III, Hammersmith Infant Neurological Examination, diagnosed cerebral palsy, gross motor function classification score, physician’s report on general development, head control, sitting, walking, and fine motor function, parents’ report of speech and walking ability, diagnoses and visual or hearing impairment. If present, the Bayley-III scores were used to define the grade of impairment as follows: normal (score ≥ −2 SD), and impaired (score < −2 SD). When Bayley-III results were partially or totally missing, all available data from the following variables at follow-up were used: Hammersmith Infant Neurological Examination, diagnosed cerebral palsy, gross motor function classification score, physician’s report on general development, head control, sitting, walking, and fine motor function, parents’ report of speech and walking ability, diagnoses and impairments of vision or hearing. The template for this grading can be found in Table S1.

### Gut microbiota

16S rRNA gene sequencing was performed on fecal samples collected weekly during the first four weeks (w) of life (1w, 2w, 3w and 4w) as previously described (27,31). Briefly, the V3-V4 hypervariable region of the 16S rRNA gene was sequenced in a paired-end 300 bp sequencing run performed in a MiSeq platform (Illumina). An amplicon sequence variant (ASV) table was generated with the DADA2 Workflow (version 1.10.1)(32).

### Statistical analyses

Background factors and clinical characteristics were analyzed and compared between groups of infants with normal versus impairment neurodevelopment, using t-test for independent samples for continuous data. Categorical data was compared with chi ^2^ – or – if any observed count was five or less, Fisher’s exact test. The background statistics were performed in IBM SPSS Statistics software, version 29 (IBM Corp, Armonk, NY, USA). The microbial causal mediation effect on language development was tested using the Sparse Microbial Causal Mediation Model (SparseMCMM) which is specific for microbiome data, as well as the Casual Mediation Analysis (*mediation* package version 4.5.0) to test for the alpha-diversity mediation (33). Alpha-diversity was calculated using Shannon’s diversity index, Pielou’s evenness index, and richness assessed as number of observed ASVs, using the *mia* package (version 1.7.5), and statistically tested for differences between the groups (normal vs impairment, and *L. reuteri* vs placebo) using Student’s t-Test. The p-values for the alpha-diversity analysis were corrected for false discovery rate according to Benjamini & Hochberg (q-value). Inference of differential abundance between the study groups was performed at ASV level and at genus taxonomic level using the Analysis of Compositions of Microbiomes with Bias Correction (ANCOM-BC) (34), with a prevalence taxa threshold of ≥ 10% (35). Prior to beta-diversity analyses, variance stabilizing transformation (VST) was applied for normalization across samples (36), using the *DESeq2* package (37). Bacterial community distributions across the normal and impairment groups were displayed by non-metric multidimensional scaling (NMDS) plots, and statistically tested using the analysis of similarities (ANOSIM), with 999 permutations (24, 32). Partial Least Squares – Discriminant Analysis (PLS-DA) was used as a supervised method to model the community at genus and ASV-level for the identification of signatures related to each outcome, using the *mixOmics* package (39). The identification of a microbial signature over time that discriminates between the study groups was performed using the *coda4microbiome* R package (40). For that, only individuals with at least three observations across the four weeks (Table 1) and taxa with a prevalence threshold ≥ 10% were used. Statistical analyses were performed in R version 4.2.1.

**Table 1.** Number of infants at each group at 1 week (w), 2w, 3w, and 4w of life. NDI: Neurodevelopment impairment. Longitudinal: number of infants with at least three observations across the four weeks.

|  |  | 1w | 2w | 3w | 4w | Longitudinal |
| --- | --- | --- | --- | --- | --- | --- |
| Language | Normal | 36 | 38 | 35 | 34 | 35 |
|  | Impairment | 31 | 29 | 31 | 27 | 30 |
| Cognition | Normal | 49 | 48 | 46 | 46 | 46 |
|  | Impairment | 22 | 22 | 23 | 19 | 22 |
| Motor | Normal | 49 | 49 | 46 | 45 | 47 |
|  | Impairment | 13 | 14 | 15 | 13 | 13 |
| NDI | Normal | 48 | 50 | 43 | 44 | 47 |
|  | Impairment | 42 | 42 | 45 | 41 | 41 |

## RESULTS

### EPT-ELBW infant cohort

From 134 infants originally enrolled in the PROPEL-trial, a two-year follow-up was only possible on 110 infants, from which, we had 16S amplicon data from 105 infants (Table 1). Among the background characteristics of the infants (Table 2 and Table S2), several clinical characteristics generally significantly differed between the normal and impairment group, including: gestational age, birth weight, birth length, birth head circumference, bronchopulmonary dysplasia (BPD), Apgar at 10 minutes and days on antibiotics. Consistently, the infants in the impairment group had the more adverse outcome of the above-mentioned clinical characteristics, i.e. infants were born at earlier gestational weeks, were smaller, were more days on antibiotics, and had more prevalence of BPD and smokers in the family.

**Table 2.** Background and clinical characteristics of the extremely preterm – extremely low birth weight infants from which stool samples collected at week 1 of life were compared to Language outcome at 2.

| Variables | week 1 |  |  |
| --- | --- | --- | --- |
|  | Normal (n=36) | Impairment (n=31) | p-value<sup>a</sup> |
| Gestational age, weeks, mean (SD) | 25.7 (1.3) | 25.2 (1.3) | 0.08 |
| Birth weight, g, mean (SD) | 790 (123) | 682 (123) | **0.001** |
| Birth weight z-score, mean (SD) | -0.8 (1.3) | -1.3 (1.3) | 0.10 |
| Birth length, cm, mean (SD) | 33.5 (2.2) | 31.8 (2.4) | **0.01** |
| Birth length, z-score, mean (SD) | -1.0 (1.7) | -1.7 (1.7) | 0.10 |
| Birth head circumference, cm, mean (SD) | 23.4 (1.4) | 22.5 (1.5) | **<0.001** |
| Birth head circumference z-score, mean (SD) | -0.7 (0.7) | -0.8 (0.7) | 0.40 |
| Apgar at 10 minutes, mean (SD) | 8.3 (2) | 7.5 (2) | 0.07 |
| SGA, n (%) | 28 (78) | 23 (74) | 0.78 |
| Female sex, n (%) | 17 (47) | 17 (55) | 0.53 |
| Infants from multiple pregnancy, n (%) | 14 (39) | 8 (26) | 0.26 |
| Prenatal steroids administered, n (%) | 35 (97) | 31 (100) | 1.00 |
| Chorioamnionitis, n (%) | 9 (25) | 6 (19) | 0.58 |
| Cesarean section, n (%) | 25 (69) | 19 (61) | 0.48 |
| Smoking in family at inclusion, n (%) | 0 (0) | 5 (16) | **0.02** |
| Inclusion site |  |  |  |
| Linköping, n (%) | 10 (28) | 18 (58) | **0.02** |
| Stockholm, n (%) | 26 (72) | 13 (42) |  |
| **Neonatal hospital care** |  |  |  |
| Bronchopulmonary dysplasia, n (%) | 19 (53) | 24 (77) | **0.04** |
| Sepsis, n (%) | 7 (19) | 10 (32) | 0.23 |
| Retinopathy of prematurity, grade 3-5, n (%) | 4 (11) | 9 (29) | 0.06 |
| Intraventricular haemorrhage, grade 3-4, n (%) | 3 (8) | 3 (10) | 1.00 |
| Necrotizing enterocolitis, grade 2-3, n (%) | 1 (3) | 2 (6) | 0.44 |
| Days on antibiotics, mean (SD) | 21 (12) | 33 (15) | **<0.001** |

<sup>a</sup>t-test for independent samples for continuous data. $\chi^2$ -test for categorical data (Fisher's exact test when appropriate).

### *L. reuteri* supplementation in relation to neurodevelopment outcomes

For this reduced nested cohort, from the PROPEL study, with matched data available for the microbiota composition and Bayley-III test, *L. reuteri* supplementation during the first three weeks of life was associated with improved language score at 24 months corrected age (p-value = 0.04, q-value=0.06, Table S3). *L. reuteri* supplementation was also significantly associated to alpha-diversity (higher alpha-diversity in the supplemented group), and beta-diversity (Table S3). *Lactobacillus* was the only taxa found to significantly differ between both groups, being more abundant in the *L. reuteri* group (Table S3). Therefore, in this study we sought to assess whether the observed effect of *L. reuteri* supplementation and language scores was mediated by the gut microbiota by assessing the mediation effect of the microbial taxa as well as the alpha-diversity. However, we could not observe any effect of the microbiota mediating the association between *L. reuteri* DSM 17938 supplementation and neurodevelopment outcomes (Table S4-S5). Given the lack of significant associations between the *L. reuteri*/placebo groups and cognition development score, motor development score, overall neurodevelopment, we proceeded to the investigation of potential associations between the gut microbiota and neurodevelopment outcomes without taking into consideration whether the infants received *L. reuteri* or placebo supplementation. *L. reuteri* supplementation was included as a confounding factor for the language outcome.

### Gut microbiota dominated by *Staphylococcus*

The gut microbiota of EPT-ELBW infants during the first month of life was dominated by few genera *Staphylococcus, Enterococcus, E. coli, Klebsiella,* and *Lactobacillus*, respectively representing in average 26%, 14%, 14%, 11% and 9%, of the total microbial community abundance.

The normal and impaired language development groups were clearly dominated by *Staphylococcus, Lactobacillus, Enterococcus,* and *E. coli* during first month of life (Figure 1A). These four genera together represented about 86%, 79%, 70%, and 65% of the total microbial community relative abundance at 1w, 2w, 3w, and 4w, respectively. These dominant genera were mostly equally represented in both groups, except for *Staphylococcus* and *Lactobacillus* at 1w, where the normal group had slightly higher relative abundance of *Staphylococcus* and less of *Lactobacillus*. At 4w, *Staphylococcus* (23%)*, E. coli* (18%)*, Enterococcus* (18%), and *Lactobacillus* (5%) covered 65 % of the total relative abundance at the impaired group, but the normal group was dominated by *Staphylococcus* (28%)*, Enterococcus* (10%)*, Klebsiella* (10%), and *Veillonella* (8%), although *Lactobacillus* also represented 5% of the relative abundance. As observed for language, the bacterial community during the first three weeks of life, for normal cognition and motor development, was dominated by *Staphylococcus, Lactobacillus, Enterococcus,* and *E. coli,* together representing in average between 73% and 87% of total microbial community abundance (Figure 1B-C). At 4w, *Lactobacillus* was no longer among the dominant bacteria, instead *Klebsiella, Enterobacter, Veillonella,* and *Bacteorides*, gained representation.

**Figure 1.**
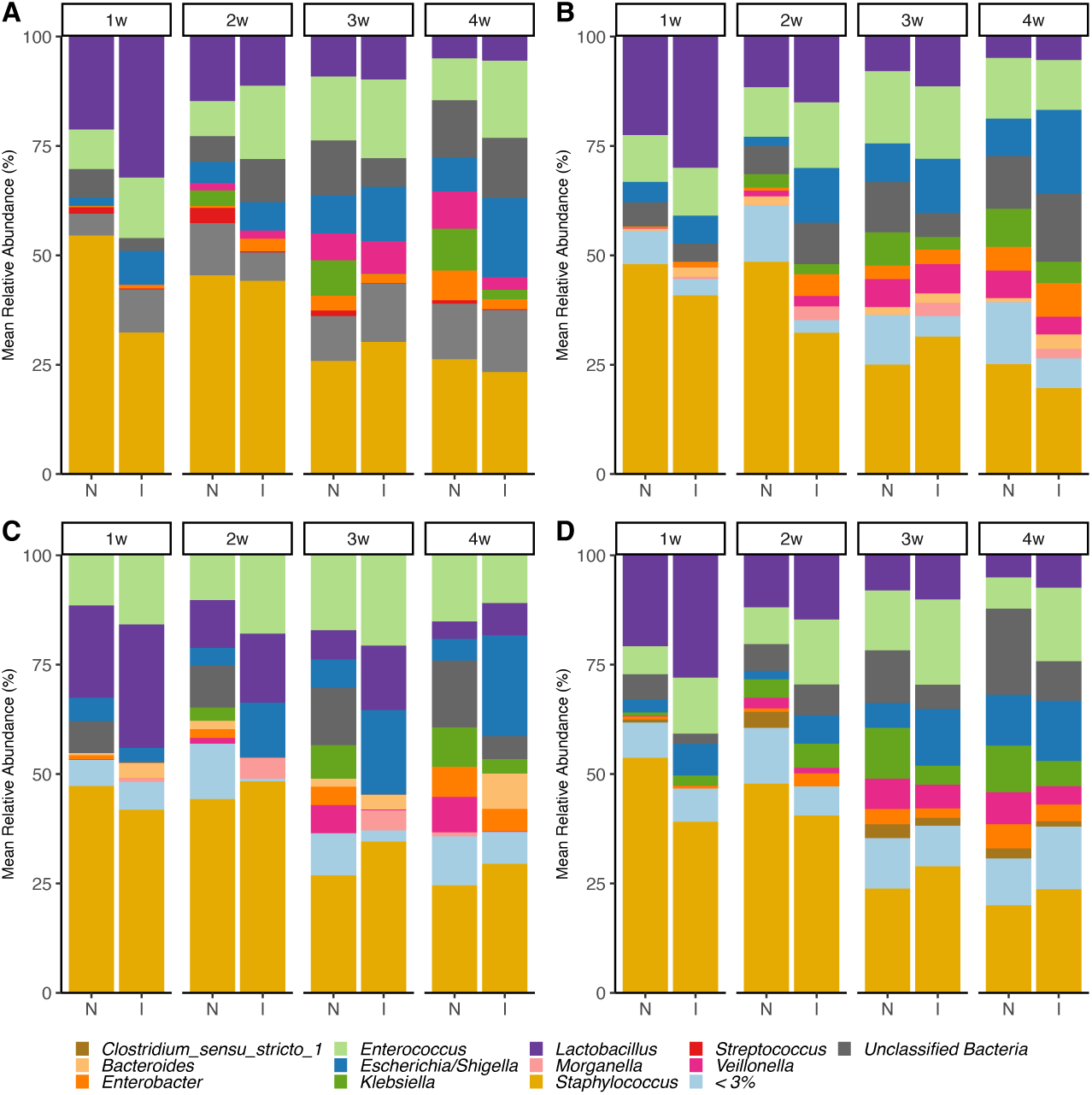
Taxonomic composition, at genus level, of the gut microbiota in ELBW-EPT infants with normal (N) *vs* impaired (I) language development (A), cognition development (B), motor development (C), and overall neurodevelopment (D) at 2 years of age. < 3% includes all genera with a relative abundance of < 3% across all samples within each data set.

Similar to the other Bayley datasets, the microbial community for the overall neurodevelopment impairment (NDI) group was dominated by *Staphylococcus, Lactobacillus,* and *Enterococcus* during the first two weeks of life, representing between 68% and 81% of the bacterial community (Figure 1D). At 3w, 50% of the relative abundance was covered by *Staphylococcus, Enterococcus* and *Klebsiella,* while at 4w the composition was more evenly distributed.

However, no significant differential abundant bacteria (at genus or ASV level at a prevalence taxa threshold of ≥ 10%) were identified between the normal and impaired groups for each timepoint, as assessed with ANCOM-BC, adjusting for the potential confounding factors (Table 2). Nonetheless, in some cases for cognition and motor development certain genera were exclusively found in the normal group (Table 3). We further explored whether there was a difference in Bayley-III scores associated to colonization of these bacteria only present in the normal development group, and we found that infants colonized with *Finegoldia* at 3w had higher Bayley-III score for cognition (p-value = 0.02, q-value = 0.14).

**Table 3.**
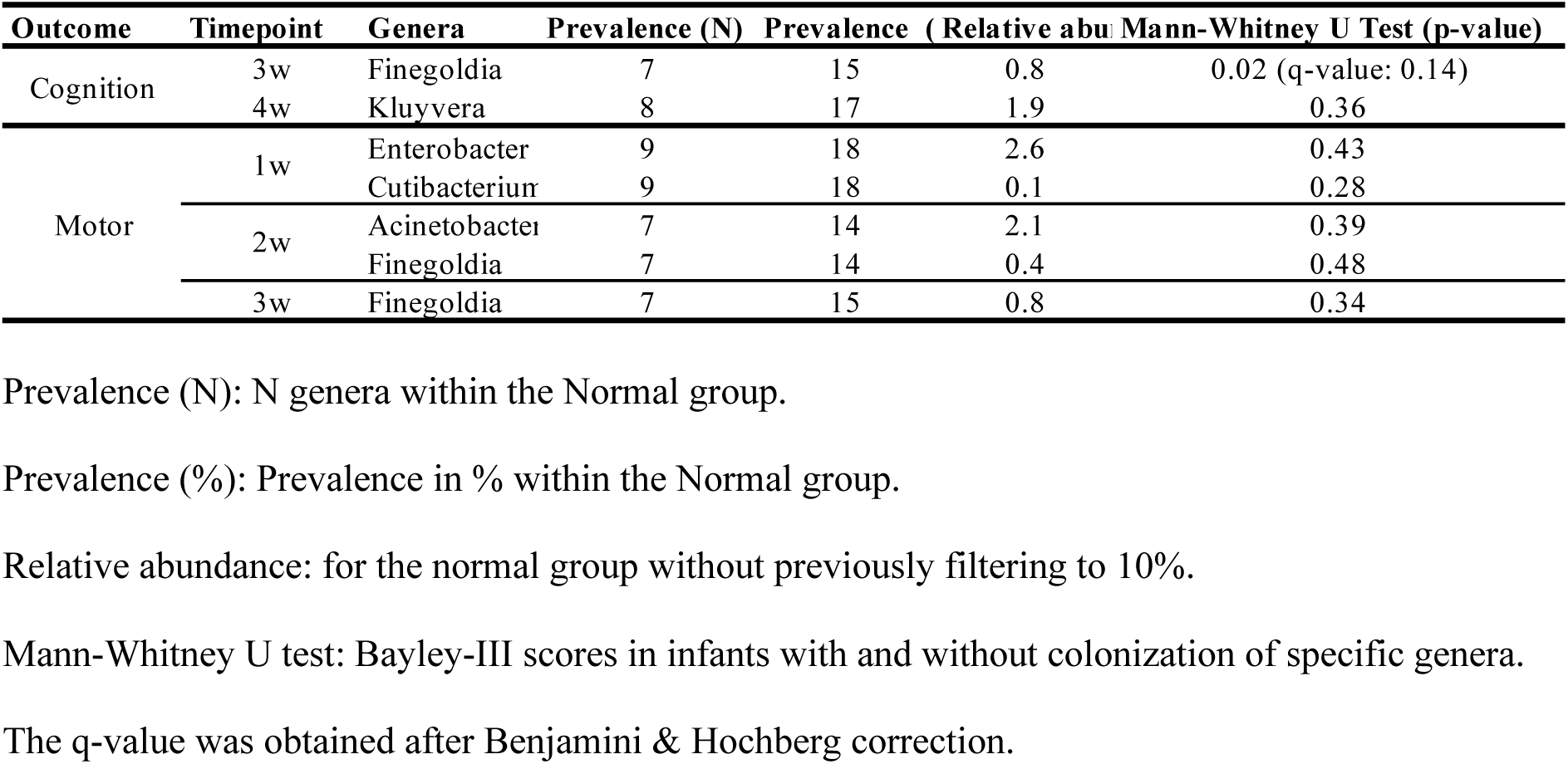
Prevalence and relative abundance of genera only present at the normal group in respect to the impaired group.

### Higher microbial richness at 3 weeks of life associated with language development

We also assessed whether the neurodevelopment was associated with differences in the community structure (alpha-diversity). At 1w, we corrected for the significant association between richness and location. For language development, less richness, less diversity, and less evenness at 3w of age, were significantly associated with language impairment (Figure 2A; q-value = 0.034). The same tendency was observed for diversity and evenness at 2w and 4w, while richness was consistently lower in the impairment group throughout the first month of life. When stratifying the language outcome by *L. reuteri* supplementation, less richness, less diversity, and less evenness was significantly associated with impairment at 3w of age for the placebo group (FigureS1). The overall NDI had the same community structure profile as language development (Figure 2D), with significant less richness in the impaired group at 2w (p-value=0.03, q-value=0.09) and 3w (p-value=0.04, q-value=0.07). For cognition and motor development, no significant differences were found, except for the stochastic significance (p-value=0.01, q-value=0.049) of less gut microbiota richness at 1w being associated with motor impairment. Nonetheless, as observed for language, there was a tendency towards lower diversity during the first month of life in the impaired groups, except for diversity and evenness and 1w (Figure 2).

**Figure 2.**
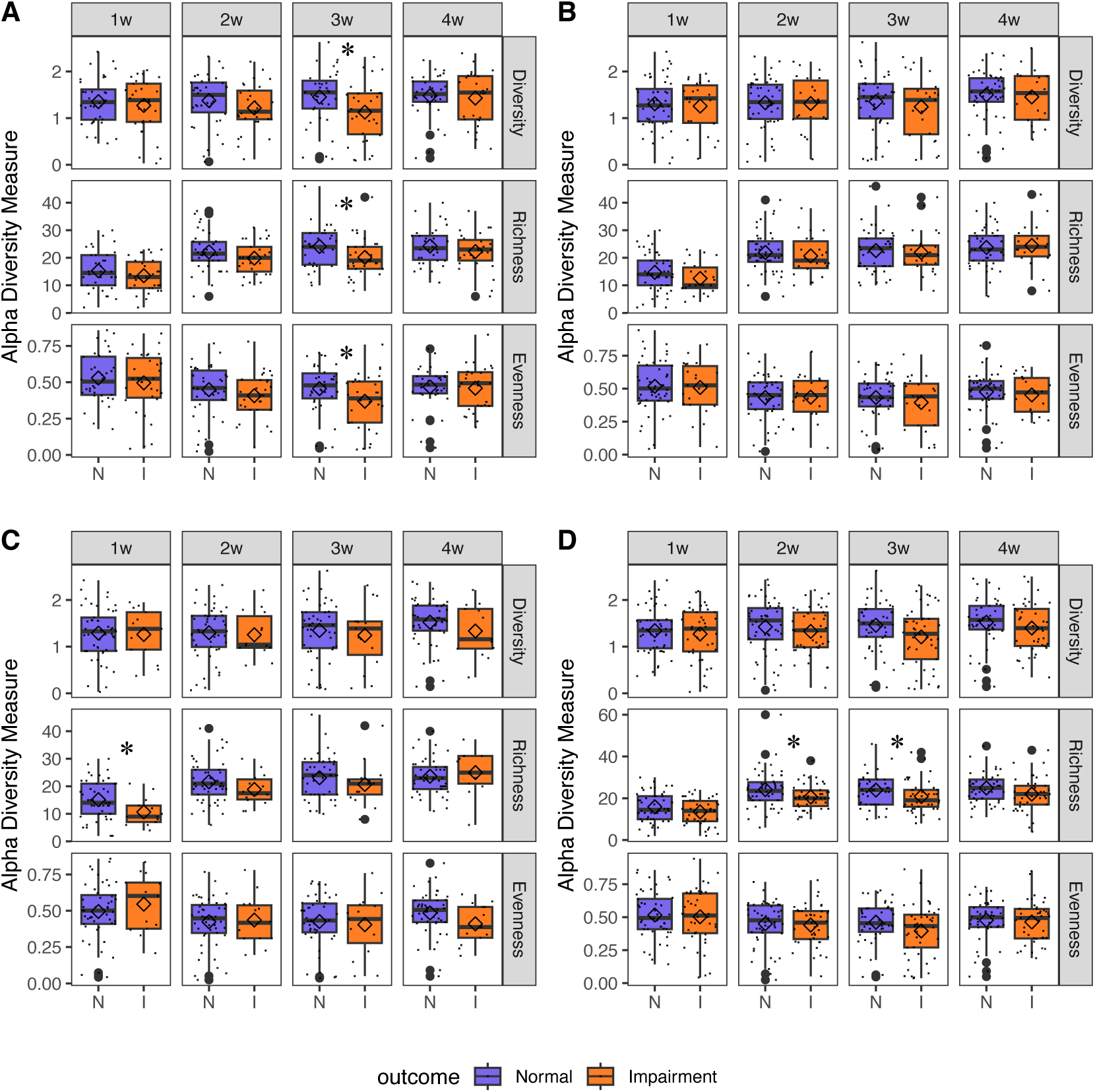
Gut microbiota alpha-diversity of ELBW-EPT infants with normal (N) or impaired (I) language development (A) cognition development (B), motor development (C), and overall neurodevelopment (D) at 2 years of age.

Boxplots (median with 25% and 75% percentiles and 1.5x the interquartile range; diamond shape depicts the mean) showing the a-diversity (Shannon index), richness (observed ASVs), and evenness (Pielou’s evenness index) from 1 week to 4 weeks of life. Student’s t-Test.

### Distinct microbial community composition between normal and impaired outcomes

Differences in beta-diversity, at ASV-level, between the normal and impaired groups at each timepoint were assessed with NMDS and ANOSIM, revealing no significant difference in the microbial community composition between the groups (Figure S2). To increase the discrimination between the normal and impaired groups, a supervised PLS-DA was performed at ASV-level, which reveled distinct microbial community compositions among the normal and impaired groups for each timepoint (Figure 3). Because the separations were mainly driven by component 1, we selected the main ASVs contributing to component 1 based on their loading weight, in order to characterize the ASVs signature associated to each group (Table S6). However, the most important ASVs contributing to both groups were assigned to *Staphylococcus sp., Lactobacillus sp*., and other taxa among the dominant ones. A PLS-DA at genus-level did not reveal any clear separation between the two groups for any outcome, thus confirming the lack of identification of key genera discriminative of normal or impaired language, cognition, motor, and overall, NDI (data not shown). The same results were found for language after stratifying by supplementation (data not shown). These results suggest it may be several bacteria in conjunction that are important for the neurodevelopment instead of individual taxa.

**Figure 3.**
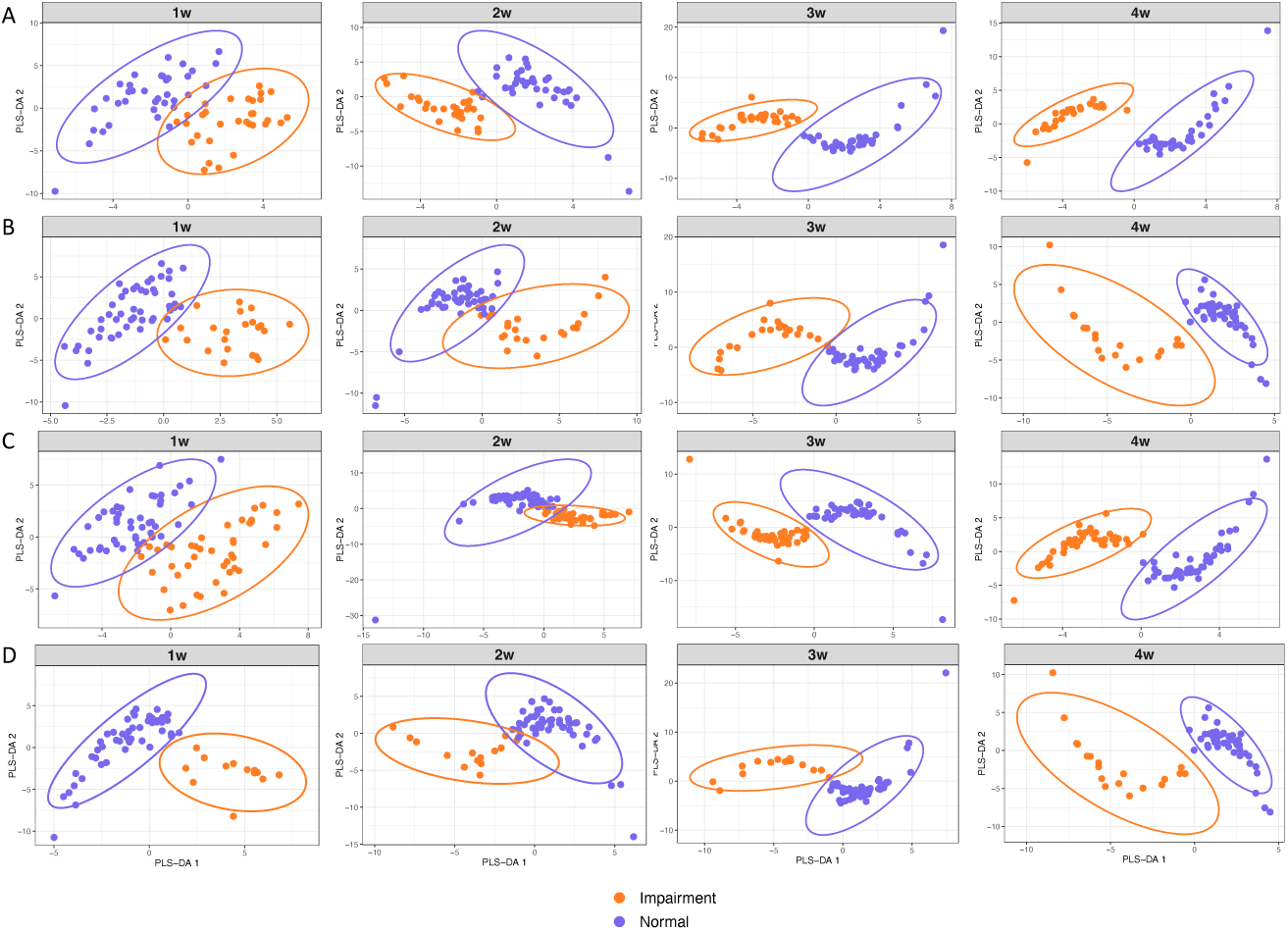
Partial least-square-discriminant analysis (PLS-DA) at ASV-level of sample distribution for language development (A), cognition development (B), motor development (C), and overall neurodevelopment (D).

### Microbiota maturation associated with neurodevelopment

The development of the gut microbiota over the first month of life was analyzed using the *coda4microbiome* to model how the development dynamics of the microbiota differed between the two groups, and to identify the group of taxa that best discriminates between the two groups. For each development outcome, we identified a microbial signature whose trajectories over the first month life differed between the normal and impaired group (Figure 4). A microbial signature consists of the relative abundances of two groups of taxa, one associated to normal development (GroupN) and the other to impaired development (GroupI), and the trajectories of the microbial signature over time depict the abundance of a group relative to the abundance of the other group. Based on the accuracy measures, the model for motor development was the best one (mean cross-validation (cv) AUC = 0.75) in contrast to the cognition development model which had a mean cv-AUC of only 0.49 (Table S7).

**Figure 4.**
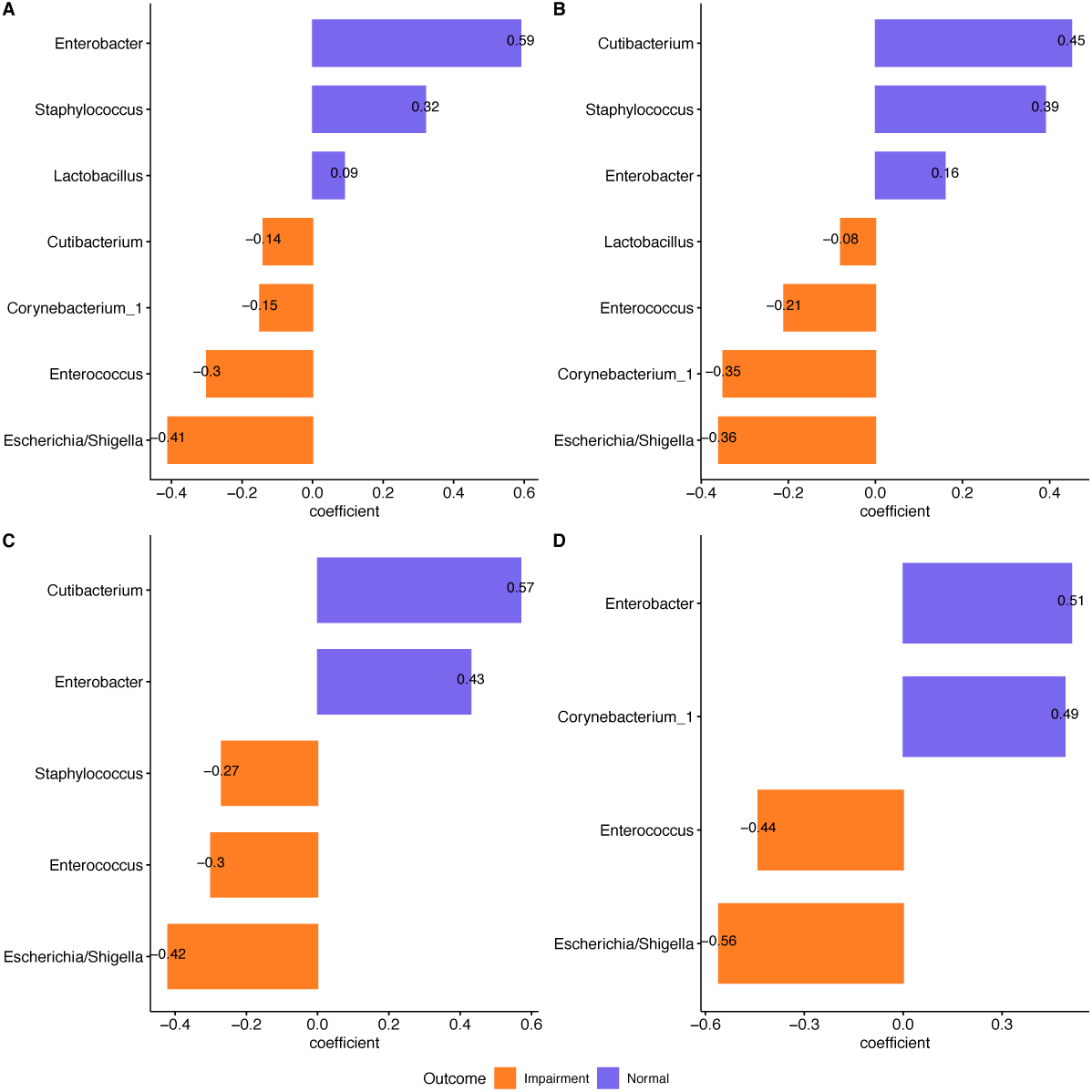
Microbial signature that discriminates between the normal and impaired groups for language development (A), cognition development (B), motor development (C), and overall neurodevelopment (D). The coefficients represent the contribution of each taxa to the model. The microbial signature was identified using the function coda_glmnet_longitudinal().

The microbial signature for motor development consisted of *Cutibacterium* and *Enterobacter* (GroupN), and *E. coli, Enterococcus,* and *Staphylococcus* (GroupI; Figure 4C). Infants with impaired motor development had higher mean relative abundance of *E. coli, Enterococcus,* and *Staphylococcus* in relation to the relative abundance of the taxa in GroupN. In contrast, infants with normal motor development had higher mean relative abundance of *Cutibacterium* and *Enterobacter.* The differences in relative abundance between the taxa in each group (GroupN and GroupI) increased at w2 due to an increase in relative abundance of the taxa in GroupI with respect to GroupN, which was stable over the month (Figure 5).

**Figure 5.**
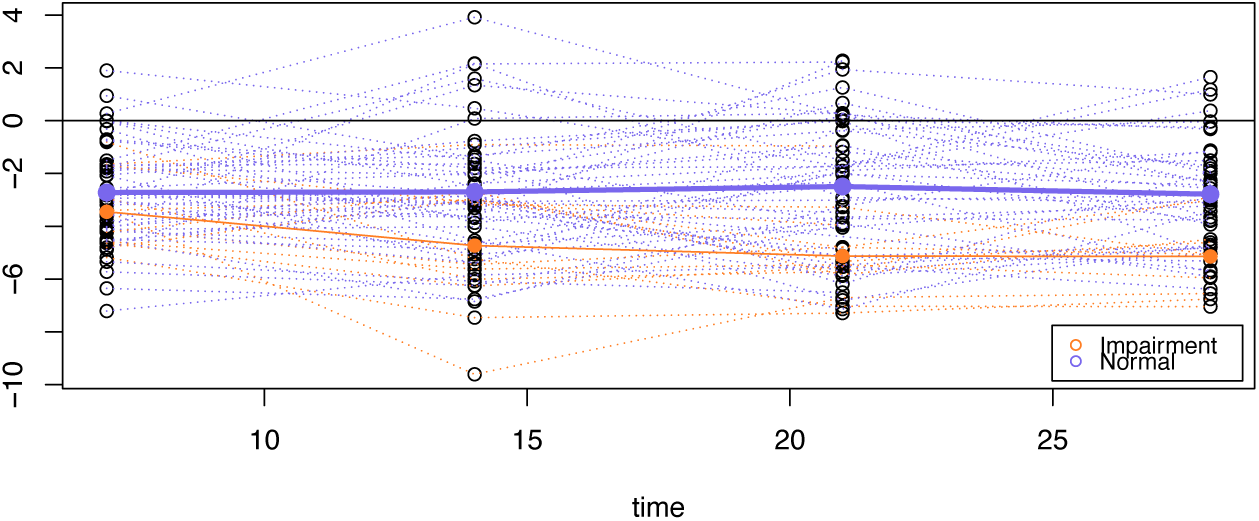
Microbial signature trajectories depicting the mean relative abundance of the microbial signature discriminative for normal and impairment group for motor development. X-axis time denotes 1w, 2w, 3w, and 4w.

The microbial signature of the infants with normal language development was composed of *Enterobacter*, *Staphylococcus*, and *Lactobacillus* (GroupN), although *Lactobacillus* contributed very little to the signature (Figure 4A). The microbial signature of the infants with language impairment was defined by *Escherichia/Shigella*, *Enterococcus*, *Corynebacterium*, and *Cutibacterium* (GroupI), with *Escherichia/Shigella* and *Enterococcus* being the main contributors. The infants with a normal language development had higher, and stable, relative abundance of *Enterobacter*, *Staphylococcus,* and *Lactobacillus* relative to the abundances of the taxa in GroupI (Figure S3). In contrast, the infants with language impairment had higher relative abundance of *Escherichia/Shigella*, *Enterococcus*, *Corynebacterium*, and *Cutibacterium* which further increased at w3 and w4.

For cognition the main taxa contributing to the GroupN were *Cutibacterium and Staphylococcus*, and the main taxa contributing to the GroupI were *Escherichia/Shigella* and *Corynebacterium* (Figure 4B). The microbial signature for NDI consisted of *Enterobacter* and *Corynebacterium* (GroupN) *vs Escherichia/Shigella* and *Enterococcus* (GroupI), (Figure 4D).

## DISCUSSION

The results of this study could not establish gut microbiota as mediating the effect of *L. reuteri* DSM 17938 supplementation on neurodevelopment. Yet, we show that rather than individual bacteria, it is specific microbiota compositions as indicated by alpha and beta diversity, and maturation during the first month of life that are associated with neurodevelopment.

The fact that we could not identify any specific taxa to be discriminative between EPT-ELBW infants with normal *vs* impaired neurodevelopment could be explained by the unique microbiota composition of these infants, which is constrained by the distinct conditions of preterm infants, including, among other, antibiotic treatment, postmenstrual age, delivery mode, delayed enteral feeds, breast milk, and NICU stay (12,18,41–43). Indeed, the taxonomic profile in our dataset is characteristic of the preterm infants during early life, which is typically dominated by *Staphylococcous, Entetrococcus, E. coli,* and *Klebsiella,* all potential pathobionts, and *Lactobacillus* (18,20–22,27,41,44). Due to the proactive care of EPT-ELBW infants in Sweden our cohort consisted of more immature infants than the other cohorts assessing the relationship between gut microbiome and neurodevelopment (45–47). The rate of antibiotic treatment was very high during the first 4 weeks of life (48), which may explain why our study did not confirm the association between low abundance of *Bifidobacterium* during the neonatal period and neurodevelopmental impairment reported in previous trials (45,46), despite exclusive feeding with breast milk due to donor milk banks during the neonatal period. Another explanation for the lack of significant individual taxa in discriminating between both groups, may be that the low gestational age and other exposures such as hypoxia, hypotension, hyperglycemia and poor nutrition of the EPT infants may be more important than an individual bacterium. However, the low microbial diversity in EPT-ELBW infants with subsequent neurodevelopmental impairment suggests that the overall composition of the gut microbiome is a contributing factor.

Seki et *al.* has reported an increase in absolute abundance of *Klebsiella* at 4 weeks of age in EPT infants to be associated with brain damage severity assessed with magnetic resonance imaging (MRI) during the neonatal period (22). Assessing actual clinical outcome later in infancy using the Bayley tool, rather than using radiologic findings, is a strength in this study. A limitation is the early assessment at 2 years of age, since an assessment at school age may be more accurate. Additionally, the methodology and data analysis *per se* may also be a source of discrepancies in the results. In our study we applied the ANCOM-BC to assess taxa that may be differently abundant between groups (34), while Seki et *al.* (22) used a completely different approach. Another explanation for the lack significant individual taxa in discriminating between both groups, may be that the physiological problems of the EPT infants may be more important than an individual bacterium.

In adults, gut microbiota alpha-diversity is generally positively associated to health (49,50) and a low diversity has been related to psychiatric disorders (51). Accordingly, in this study we found that the EPT-ELBW infants with normal language development as well as a normal overall neurodevelopment (NDI) had higher alpha-diversity throughout the first month of life. However, other studies on term infants have reported higher diversity to be related to adverse neurocognitive outcomes. For example, Carlson 2018 (16) showed that diversity at 1 year of age was negatively associated with cognitive performance at 2 years of age, Loughman 2020 (52) found slightly higher diversity in infants with behavior problems at 1 year of age, and Chen 2022 (15) reported that term-infants small for gestational age had with poor communication scores at 6 months of age, higher diversity at three days of life. Nonetheless, all these studies differ in the timepoints when diversity and neurocognitive outcomes are associated. Differences in sequencing technologies and data analyses may also be a source for discrepancies (53). However, it is worth noticing that term-infants have higher diversity than preterm infants (20,21), and thus, we could speculate that the higher diversity in our cohort may resemble the lower diversity in term infants. This would indicate that there may be certain gut microbiota constellations beneficial for the neurodevelopment in infants which in turn may be specific to infant cohort. Indeed, the distinct gut microbiota composition observed between the normal and impairment groups supports this hypothesis.

The importance of the composition of the gut microbiota community was further sustained by the longitudinal analysis which revealed, for each development outcome, a microbial signature whose trajectories differed over the first month of life. The development of the gut microbiota composition and function from early-life has been shown to be dynamic and may stabilize between 3 to 5 years of age (41,54–56). For term infants, the development phases start with and immature *E.coli-*dominated community to mature towards a *Bacteroides*-dominated community (55). For preterm infants, Korpela et al. (18) described four phases of microbiota development from birth up to 60 days postnatal age, with a transition from *Staphylococcus*-dominated community, to *Enterococcus-*dominated, to then *Enterobacter-*dominated, and finally *Bifidobacterium-*dominated. Our study time span falls in the first phase (birth to 35 weeks postmenstrual age) and accordingly, the gut microbiota during the four first weeks of life was a *Staphylococcus*-dominated community. *E. coli* was the bacterium contributing the most to the impairment signatures for all the four development outcomes. In our study, the main taxa contributing to the microbial signatures for all the development outcomes were *E. coli* and *Enterococcus* vs *Enterobacter,* with *E. coli* and *Enterococcus* having a higher relative abundance with respect to *Enterobacter* in the impairment group, and vice versa for the normal group. These findings are supported by previous trial reporting an association between high *Enterococcus* levels in EPT infants during the neonatal period and poor neurodevelopmental outcome at 2 years of age (46,47). Interestingly, the taxa discriminant for the impaired group belong to early maturation stages in the gut microbiota development, while *Enterobacter* is representative of a more mature stage (41,55). Given this observation, our results suggest that a more mature microbiota composition already during the first month of life may have positive consequences on neurodevelopment. This is further supported by the trajectories in the motor outcome, the one with the highest discrimination accuracy (AUC = 0.75), where the impairment group besides having higher relative abundance of *E. coli* and *Enterococcus,* those abundances further increased from week 1 to week 4.

Although daily supplementation with *L.reuteri DSM 17938* during the neonatal period (until postmenstrual week 36+0) was associated with improved language score at 24 months corrected age in this cohort (29), we could not identify a mediation mechanism via the gut microbiome. Therefore, we hypothesize that *L.reuteri* may exert a direct effect on the central nervous system (57), or an indirect effect thought an unidentified factor.

In conclusion, this prospective observational study indicates that rather than individual taxa, it is alpha diversity, a balance between certain taxa and, importantly, its maturation over the first month of life that may have an impact on the neurodevelopment at 24 months.

## Data Availability

All data produced in the present study are available upon reasonable request to the authors

## ACKNOWLEDGEMENTS

We thank Professor M. Luz Calle and Msc. Meritxell Pujolassos for their support on the application of the coda4microbiome computational analysis. The computations were partially performed on resources provided by the Swedish National Infrastructure for Computing (SNIC) through Uppsala Multidisciplinary Center for Advanced Computational Science (UPPMAX) under project SNIC 2020/5-336. We also thank Dr Fredrik Ingemansson, Dr Josefin Lundström, Dr Anders Palm, Dr Björn Westrup and, Dr Laura Österdahl and the study nurses Mrs Christina Fuxin and Mrs Karin Jansmark for their help with the trial; and Dr Stellan Håkansson for help with the SNQ database.

## DATA AVAILABILITY STATEMENT

The 16S rRNA dataset generated in this study is available at the European Nucleotide Archive: PRJEB36531.

