## Supplemental Material for "Dynamics of early gut microbiota maturation in extremely preterm infants in relation with neurodevelopment at 2 years"

**Supplementary Table S1.** Form for assessing neurodevlopmental impairment (NDI) using Bayley-III and/or clinical data.

| Developmental domain | Classification of neuroimpairment in each developmental domain | | | |
|---|---|---|---|---|
| | **None (0)** | **Mild (1)** | **Moderate (2)** | **Severe (3)** |
| **Cognition OR language** | | | | |
| Bayley III cognitive scales | Bayley III Scores > -1 SD (Index ≥ 95) | Bayley III Scores -1 SD to -2 SD (Index 83-94) | Bayley III Scores -2 SD to -3 SD (Index 72-82) | Bayley III Scores < -3 SD (Index <72) |
| Bayley III language scales | Bayley III Scores > -1 SD (Index ≥ 97) | Bayley III Scores -1 SD to -2 SD (Index 85-96 | Bayley III Scores -2 SD to -3 SD (Index 72-84) | Bayley III Scores < -3 SD (Index <72) |
| Parent report | Speech: Uses sentences with 2-3 words | Speech: Says a few words (vocabulary > 10 words) | Speech: Says a few words (vocabulary < 10 words) | Speech: Does not speak at all |
| **OR Motor function** | | | | |
| CP (GMFCSF) | None | 1 | 2-3 | 4-5 (non ambulant) |
| Bayley III fine and gross motor scales | > -1 SD (Index ≥ 94) | Scores -1 SD to -2 SD (Index 80-93) | Scores -2 SD to -3 SD (Index 66-79) | Scores < -3 SD (Index <66) |
| Doctor´s report | Normal head control<br>Sits without support<br>Walks without support<br><br>Normal finger movements | Fine motor function: Clumsy but can grasp<br>OR<br>Abnormal neurology or motor development | Fine motor function: Clumsy but can grasp | Unsteady head control<br>Unsteady sitting or cannot sit.<br>Fine motor: Does not grasp objects. |
| **OR Hearing** | | | | |
| Parent report | No hearing impairment | No hearing impairment | Hearing impairment<br>OR<br>Has hearing aids but still impaired hearing | Cannot hear |
| **OR Vision** | | | | |
| Parent report | No visual impairment | Visual impairment: No impairment when wearing glasses | Visual impairment: Serious visual impairment that remains despite glasses<br>OR<br>Admitted to centre for visual impairments | Visual impairment: Bilateral blindness |

**Supplementary Table S2.** Background and clinical characteristics of the extremely preterm – extremely low birth weight infants from which samples collected at week 1, week 3, week3, and week 4 of life and were compared to Language, Cognition, Motor, and NDI outomes s at 2 years of ages.

| | | | week 1 | | | week 2 | | | week 3 | | | week 4 | | |
|---|---|---|---|---|---|---|---|---|---|---|---|---|---|---|
| | **Variables** | **Statistical test** | **Normal (n=36)** | **Impairment (n=31)** | **p value** | **Normal (n=38)** | **Impairment (n=29)** | **p value** | **Normal (n=35)** | **Impairment (n=31)** | **p value** | **Normal (n=34)** | **Impairment (n=27)** | **p value** |
| **Language** | Gestational age, weeks, mean (SD) | t-test for independent samples | 25.7 (1.3) | 25.2 (1.3) | 0.08 | 25.8 (1,0) | 25.2 (1.4) | **0.03** | 25.8 (1) | 25.2 (1.3) | 0.05 | 25.7 (1) | 25.2 (1.3) | 0.10 |
| | Birth weight, g, mean (SD) | t-test for independent samples | 790 (123) | 682 (123) | **0.001** | 791 (120) | 686 (125) | **0.001** | 798 (109) | 692 (125) | **0.001** | 779 (119) | 684 (110) | **0.002** |
| | Birth weight z-score, mean (SD) | t-test for independent samples | -0.8 (1.3) | -1.3 (1.3) | 0.10 | -0.9 (1.1) | -1.3 (1.3) | 0.10 | -0.9 (1) | -1.3 (1.3) | 0.17 | -0.9 (1.1) | -1.3 (1.1) | 0.24 |
| | Birth length, cm, mean (SD) | t-test for independent samples | 33.5 (2.2) | 31.8 (2.4) | **0.01** | 33.4 (2.3) | 32 (2.4) | **0.02** | 33.5 (2.2) | 32.1 (2.4) | **0.01** | 33.3 (2.3) | 32.1 (2.3) | **0.04** |
| | Birth length, z-score, mean (SD) | t-test for independent samples | -1,0 (1.7) | -1.7 (1.7) | 0.10 | -1.2 (1.8) | -1.6 (1.7) | 0.19 | -1.1 (1.6) | -1.5 (1.6) | 0.26 | -1.1 (1.7) | -1.5 (1.5) | 0.42 |
| | Birth head circumference, cm, mean (SD) | t-test for independent samples | 23.4 (1.4) | 22.5 (1.5) | **<0,001** | 23.5 (1.4) | 22.6 (1.5) | **0.01** | 23,6 (1.4) | 22.6 (1.5) | **0.01** | 23.5 (1.5) | 22.6 (1.4) | **0.01** |
| | Birth head circumference z-score, mean (SD) | t-test for independent samples | -0.7 (0.7) | -0.8 (0.7) | 0.40 | -0.6 (0.8) | -0.8 (0.6) | 0.26 | -0.6 (0.8) | -0.8 (0.7) | 0.42 | -0.6 (0.9) | -0.8 (0.7) | 0.29 |
| | Apgar at 10 minutes, mean (SD) | t-test for independent samples | 8.3 (2) | 7.5 (2) | 0.07 | 8.2 (1.7) | 7.6 (1.8) | 0.10 | 8.4 (1.4) | 7.6 (1.7) | **0.04** | 8.2 (1.6) | 7.1 (1.9) | **0.02** |
| | SGA, n (%) | Chi-square or Fisher's exact test as appropriate | 28 (78) | 23 (74) | 0.78 | 9 (24) | 7 (24) | 0.97 | 7 (20) | 7 (23) | 0.80 | 9 (26) | 6 (22) | 0.70 |
| | Female sex, n (%) | Chi-square or Fisher's exact test as appropriate | 17 (47) | 17 (55) | 0.53 | 18 (47) | 15 (52) | 0.72 | 17 (49) | 16 (52) | 0.81 | 14 (41) | 13 (48) | 0.59 |
| | Infants from multiple pregnancy, n (%) | Chi-square or Fisher's exact test as appropriate | 14 (39) | 8 (26) | 0.26 | 17 (45) | 9 (31) | 0.25 | 16 (46) | 10 (32) | 0.26 | 15 (44) | 9 (33) | 0.39 |
| | Prenatal steroids administered, n (%) | Chi-square or Fisher's exact test as appropriate | 35 (97) | 31 (100) | 1.00 | 37 (97) | 29 (100) | 1.00 | 34 (97) | 31 (100) | 1.00 | 34 (100) | 27 (100) | n.a. |
| | Chorioamnionitis, n (%) | Chi-square or Fisher's exact test as appropriate | 9 (25) | 6 (19) | 0.58 | 10 (26) | 6 (21) | 0.59 | 9 (26) | 7 (23) | 0.77 | 8 (24) | 6 (22) | 0.90 |
| | Cesarean section, n (%) | Chi-square or Fisher's exact test as appropriate | 25 (69) | 19 (61) | 0.48 | 27 (71) | 18 (62) | 0.44 | 27 (77) | 20 (64) | 0.26 | 24 (71) | 19 (70) | 0.99 |
| | Smoking in family at inclusion, n (%) | Chi-square or Fisher's exact test as appropriate | 0 (0) | 5 (16) | **0.02** | 0 (0) | 4 (14) | **0.03** | 0 (0) | 4 (13) | **0.04** | 0 (0) | 4 (15) | **0.03** |
| | Inclusion site | Chi-square or Fisher's exact test as appropriate | | | | | | | | | | | | |
| | Linkoping, n (%) | | 10 (28) | 18 (58) | **0.02** | 11 (29) | 17 (59) | **0.01** | 10 (29) | 17 (55) | **0.03** | 9 (26) | 16 (59) | **0.01** |
| | Stockholm, n (%) | | 26 (72) | 13 (42) | | 27 (71) | 12 (41) | | 25 (71) | 14 (45) | | 25 (74) | 11 (41) | |
| | **Neonatal hospital care** | | | | | | | | | | | | | |
| | Bronchopulmonary dysplasia, n (%) | Chi-square or Fisher's exact test as appropriate | 19 (53) | 24 (77) | **0.04** | 20 (53) | 22 (76) | 0.05 | 19 (54.3) | 24 (77.4) | **0.05** | 18 (53) | 23 (85) | **0.01** |
| | Sepsis, n (%) | Chi-square or Fisher's exact test as appropriate | 7 (19) | 10 (32) | 0.23 | 9 (24) | 9 (31) | 0.50 | 9 (25.7) | 10 (32.3) | 0.56 | 9 (26) | 5 (19) | 0.46 |
| | Retinopathy of prematurity, grade 3-5, n (%) | Chi-square or Fisher's exact test as appropriate | 4 (11) | 9 (29) | 0.06 | 4 (11) | 7 (24) | 0.19 | 4 (11.4) | 7 (22.6) | 0.19 | 4 (12) | 8 (30) | 0.11 |
| | Intraventricular haemorrhage, grade 3-4, n (%) | Chi-square or Fisher's exact test as appropriate | 3 (8) | 3 (10) | 1.00 | 4 (11) | 3 (10) | 0.65 | 4 (11.4) | 3 (9.7) | 0.57 | 3 (9) | 1 (4) | 0.62 |
| | Necrotizing enterocolitis, grade 2-3, n (%) | Chi-square or Fisher's exact test as appropriate | 1 (3) | 2 (6) | 0.44 | 1 (3) | 3 (10) | 0.31 | 0 (0) | 4 (12.9) | 0.04 | 2 (6) | 3 (11) | 0.65 |
| | Days on antibiotics, mean (SD) | t-test for independent samples | 21 (12) | 33 (15) | **<0,001** | 21(12) | 33 (15) | **<0,001** | 21 (12) | 33 (15) | **<0,001** | 22 (13) | 33 (15) | **0.004** |

| | | | week 1 | | | week 2 | | | week 3 | | | week 4 | | |
|---|---|---|---|---|---|---|---|---|---|---|---|---|---|---|
| | **Variables** | **Statistical test** | **Normal (n=49)** | **Impairment (n=22)** | **p value** | **Normal (n=48)** | **Impairment (n=22)** | **p value** | **Normal (n=46)** | **Impairment (n=23)** | **p value** | **Normal (n=46)** | **Impairment (n=19)** | **p value** |
| **Cognition** | Gestational age, weeks, mean (SD) | t-test for independent samples | 25,6 (1,1) | 25,2 (1,5) | 0.20 | 25,7 (1,1) | 25,2 (1,5) | 0.14 | 25,7 (1,1) | 25,2 (1,4) | 0.13 | 25,6 (1,1) | 25,1 (1,4) | 0.20 |
| | Birth weight, g, mean (SD) | t-test for independent samples | 755 (129,7) | 686,1 (125,6) | **0.04** | 766 (131) | 692 (122) | **0.03** | 763 (124) | 699 (124) | 0.05 | 754 (126) | 689 (103) | 0.05 |
| | Birth weight z-score, mean (SD) | t-test for independent samples | -1 (1,0) | -1,3 (1,5) | 0.44 | -1,0 (1,1) | -1,2 (1,5) | 0.48 | -1,0 (1,0) | -1,2 (1,5) | 0.54 | -1,1 (1,1) | -1,3 (1,4) | 0.53 |
| | Birth length, cm, mean (SD) | t-test for independent samples | 33 (2,5) | 31,7 (2,2) | **0.03** | 33,1 (2,4) | 31,8 (2,2) | **0.03** | 33,2 (2,3) | 31,9 (2,3) | **0.03** | 33,1 (2,4) | 31,9 (2,0) | 0.07 |
| | Birth length, z-score, mean (SD) | t-test for independent samples | -1,2 (1,6) | -1,7 (2) | 0.24 | -1,2 (1,7) | -1,7 (2) | 0.32 | -1,2 (1,5) | -1,6 (2,0) | 0.33 | -1,2 (1,5) | -1,5 (1,8) | 0.47 |
| | Birth head circumference, cm, mean (SD) | t-test for independent samples | 23,1 (1,4) | 22,5 (1,4) | 0.27 | 23,2 (1,5) | 22,7 (1,4) | 0.12 | 23,2 (1,5) | 22,8 (1,5) | 0.20 | 23,2 (1,6) | 22,7 (1,4) | 0.19 |
| | Birth head circumference z-score, mean (SD) | t-test for independent samples | -0,8 (0,8) | -0,7 (0,8) | 0.57 | -0,7 (0,8) | -0,7 (0,8) | 0.82 | -0,8 (0,8) | -0,7 (0,8) | 0.56 | -0,7 (0,8) | -0,7 (0,8) | 0.84 |
| | Apgar at 10 minutes, mean (SD) | t-test for independent samples | 8,2 (1,5) | 7,5 (2,2) | 0.12 | 8,1 (1,6) | 7,7 (2) | 0.38 | 8,2 (1,4) | 7,6 (1,9) | 0.20 | 8 (1,5) | 7,0 (2,2) | **0.04** |
| | SGA, n (%) | Chi-square or Fisher's exact test as appropriate | 12 (25) | 5 (23) | 0.87 | 12 (25) | 5 (23) | 0.84 | 11 (24) | 5 (22) | 0.84 | 12 (26) | 4 (21) | 0.67 |
| | Female sex, n (%) | Chi-square or Fisher's exact test as appropriate | 27 (55) | 8 (36) | 0.14 | 26 (54) | 8 (36) | 0.17 | 27 (59) | 8 (35) | 0.06 | 23 (50) | 5 (26) | 0.08 |
| | Infants from multiple pregnancy, n (%) | Chi-square or Fisher's exact test as appropriate | 16 (33) | 8 (36) | 0.76 | 19 (40) | 9 (41) | 0.92 | 18 (39) | 10 (44) | 0.73 | 17 (37) | 9 (47) | 0.44 |
| | Prenatal steroids administered, n (%) | Chi-square or Fisher's exact test as appropriate | 48 (98) | 22 (100) | 1.00 | 47 (98) | 22 (100) | 1.00 | 45 (98) | 23(100) | 1.00 | 46 (100) | 19 (100) | n.a. |
| | Chorioamnionitis, n (%) | Chi-square or Fisher's exact test as appropriate | 11 (22) | 4 (18) | 0.76 | 11 (23) | 5 (23) | 0.99 | 12 (26) | 5 (22) | 0.69 | 10 (22) | 4 (21) | 1.00 |
| | Cesarean section, n (%) | Chi-square or Fisher's exact test as appropriate | 32 (65) | 14 (64) | 0.89 | 33 (69) | 14 (64) | 0.67 | 34 (74) | 15 (65) | 0.45 | 31 (67) | 14 (74) | 0.62 |
| | Smoking in family at inclusion, n (%) | Chi-square or Fisher's exact test as appropriate | 4 (8) | 2 (9) | 1.00 | 3 (6) | 2 (9) | 0.65 | 3 (6) | 2 (9) | 1.00 | 3 (7) | 2 (10) | 0.63 |
| | Inclusion site | Chi-square or Fisher's exact test as appropriate | | | | | | | | | | | | |
| | Linkoping, n (%) | | 18 (37) | 11 (50) | 0.29 | 19 (40) | 10 (46) | 0.64 | 18 (39) | 10 (44) | 0.73 | 17 (37) | 9 (47) | 0.44 |
| | Stockholm, n (%) | | 31 (63) | 11 (50) | | 29 (60) | 12 (55) | | 28 (61) | 13 (56) | | 29 (63) | 10 (53) | |
| | **Neonatal hospital care** | | | | | | | | | | | | | |
| | Bronchopulmonary dysplasia, n (%) | Chi-square or Fisher's exact test as appropriate | 28 (57) | 17 (77) | 0.10 | 27 (56) | 17 (77) | 0.09 | 26 (56) | 18 (78) | 0.08 | 27 (69) | 17 (89) | **0.02** |
| | Sepsis, n (%) | Chi-square or Fisher's exact test as appropriate | 10 (20) | 8 (36) | 0.15 | 11 (23) | 8 (36) | 0.24 | 11 (24) | 9 (39) | 0.19 | 11 (24) | 4 (21) | 0.80 |
| | Retinopathy of prematurity, grade 3-5, n (%) | Chi-square or Fisher's exact test as appropriate | 7 (14) | 6 (27) | 0.20 | 6 (13) | 5 (23) | 0.30 | 6 (13) | 5 (22) | 0.49 | 6 (13) | 6 (32) | 0.16 |
| | Intraventricular haemorrhage, grade 3-4, n (%) | Chi-square or Fisher's exact test as appropriate | 3 (6) | 3 (14) | 0.37 | 4 (8) | 3 (14) | 0.67 | 4 (9) | 3 (13) | 0.68 | 3 (7) | 1 (5) | 1.00 |
| | Necrotizing enterocolitis, grade 2-3, n (%) | Chi-square or Fisher's exact test as appropriate | 1 (2) | 2 (9) | 0.23 | 1 (2) | 3 (14) | 0.09 | 0 (0) | 4 (17) | 0.01 | 2 (4) | 3 (16) | 0.14 |
| | Days on antibiotics, mean (SD) | t-test for independent samples | 24 (14) | 32 (17) | **0.03** | 24 (13) | 32 (17) | **0.03** | 24 (13) | 33 (17) | **0.02** | 25 (14) | 32 (17) | 0.08 |

| | | | week 1 | | | week 2 | | | week 3 | | | week 4 | | |
|---|---|---|---|---|---|---|---|---|---|---|---|---|---|---|
| | **Variables** | **Statistical test** | **Normal (n=49)** | **Impairment (n=13)** | **p value** | **Normal (n=49)** | **Impairment (n=14)** | **p value** | **Normal (n=46)** | **Impairment (n=15)** | **p value** | **Normal (n=45)** | **Impairment (n=13)** | **p value** |
| **Motor** | Gestational age, weeks, mean (SD) | t-test for independent samples | 25,6 (1,2) | 24,8 (1,2) | **0.02** | 25,6 (1,2) | 24,9 (1,2) | 0.06 | 25,7 (1,1) | 25,0 (1,2) | 0.06 | 25,6 (1,1) | 24,8 (1,0) | **0.02** |
| | Birth weight, g, mean (SD) | t-test for independent samples | 755 (128) | 687 (139) | 0.10 | 758 (131) | 706 (133) | 0.20 | 757 (126) | 716 (134) | 0.28 | 751 (127) | 712 (118) | 0.32 |
| | Birth weight z-score, mean (SD) | t-test for independent samples | -1,1 (1,2) | -0,9 (1,4) | 0.70 | -1,1 (1,2) | -0,9 (1,3) | 0.65 | -1,1 (1,1) | -0,9 (1,3) | 0.58 | -1,2 (1,2) | -0,7 (0,8) | 0.21 |
| | Birth length, cm, mean (SD) | t-test for independent samples | 33,1 (2,5) | 31,9 (2,4) | 0.14 | 33,1 (2,4) | 31,9 (2,3) | 0.10 | 33,2 (2,3) | 32,1 (2,3) | 0.12 | 22,1 (2,4) | 32,4 (2,0) | 0.36 |
| | Birth length, z-score, mean (SD) | t-test for independent samples | -1,2 (1,6) | -1,1 (2) | 0.75 | -1,2 (1,6) | -1,3 (2,2) | 0.88 | -1,1 (1,5) | -1,2 (2,1) | 0.91 | -1,3 (1,6) | -0,7 (1,2) | 0.25 |
| | Birth head circumference, cm, mean (SD) | t-test for independent samples | 23,1 (1,5) | 22,6 (1,3) | 0.32 | 23,2 (1,5) | 22,8 (1,3) | 0.35 | 23,2 (1,5) | 22,9 (1,4) | 0.49 | 23,3 (1,6) | 22,8 (1,4) | 0.31 |
| | Birth head circumference z-score, mean (SD) | t-test for independent samples | -0,8 (0,8) | -0,4 (0,5) | 0.08 | -0,7 (0,8) | -0,4 (0,5) | 0.17 | -0,7 (0,8) | -0,4 (0,5) | 0.10 | -0,7 (0,8) | -0,3 (0,4) | **0.02** |
| | Apgar at 10 minutes, mean (SD) | t-test for independent samples | 8,3 (1,5) | 7,1 (2,4) | 0.10 | 8,2 (1,6) | 7,4 (2,0) | 0.11 | 8,4 (1,4) | 7,3 (1,9) | **0.03** | 8,2 (1,5) | 6,5 (2,0) | **0.003** |
| | SGA, n (%) | Chi-square or Fisher's exact test as appropriate | 14 (29) | 1 (8) | 0.12 | 14 (29) | 1 (7) | 0.16 | 13 (28) | 1 (7) | 0.16 | 14 (31) | 0 (0) | **0.03** |
| | Female sex, n (%) | Chi-square or Fisher's exact test as appropriate | 26 (53) | 3 (23) | 0.07 | 25 (51) | 4 (29) | 0.14 | 25 (54) | 4 (27) | 0.06 | 22 (49) | 1 (8) | **0.01** |
| | Infants from multiple pregnancy, n (%) | Chi-square or Fisher's exact test as appropriate | 17 (35) | 4 (31) | 1.00 | 19 (39) | 6 (43) | 0.78 | 18 (39) | 7 (47) | 0.61 | 18 (40) | 5 (38) | 0.92 |
| | Prenatal steroids administered, n (%) | Chi-square or Fisher's exact test as appropriate | 48 (98) | 13 (100) | 1.00 | 48 (98) | 14 (100) | 1.00 | 45 (98) | 15 (100) | 1.00 | 45 (100) | 13 (100) | n.a. |
| | Chorioamnionitis, n (%) | Chi-square or Fisher's exact test as appropriate | 10 (20) | 2 (15) | 1.00 | 10 (20) | 4 (29) | 0.52 | 10 (22) | 4 (27) | 0.69 | 9 (20) | 2 (15) | 1.00 |
| | Cesarean section, n (%) | Chi-square or Fisher's exact test as appropriate | 32 (65) | 38 (61) | 0.21 | 33 (67) | 7 (50) | 0.23 | 33 (72) | 8 (53) | 0.22 | 31 (69) | 7 (54) | 0.34 |
| | Smoking in family at inclusion, n (%) | Chi-square or Fisher's exact test as appropriate | 3 (6) | 2 (15) | 0.28 | 2 (4) | 2 (14) | 0.17 | 2 (4) | 2 (13) | 0.25 | 2 (4) | 2 (15) | 0.21 |
| | Inclusion site | Chi-square or Fisher's exact test as appropriate | | | | | | | | | | | | |
| | Linkoping, n (%) | | 17 (35) | 10 (77) | **0.01** | 18 (37) | 9 (64) | 0.07 | 17 (37) | 9 (60) | 0.12 | 16 (36) | 8 (62) | 0.09 |
| | Stockholm, n (%) | | 32 (65) | 3 (23) | | 31 (63) | 5 (36) | | 29 (63) | 6 (40) | | 29 (64) | 5 (38) | |
| | **Neonatal hospital care** | | | | | | | | | | | | | |
| | Bronchopulmonary dysplasia, n (%) | Chi-square or Fisher's exact test as appropriate | 27 (55) | 11 (85) | 0.05 | 27 (55) | 11 (79) | 0.11 | 25 (54) | 12 (80) | 0.13 | 25 (56) | 12 (92) | **0.02** |
| | Sepsis, n (%) | Chi-square or Fisher's exact test as appropriate | 10 (20) | 4 (31) | 0.47 | 10 (20) | 5 (36) | 0.29 | 10 (22) | 6 (40) | 0.19 | 10 (22) | 2 (15) | 0.72 |
| | Retinopathy of prematurity, grade 3-5, n (%) | Chi-square or Fisher's exact test as appropriate | 6 (12) | 4 (31) | 0.20 | 5 (10) | 3 (21) | 0.36 | 5 (11) | 3 (20) | 0.39 | 5 (11) | 4 (31) | 0.10 |
| | Intraventricular haemorrhage, grade 3-4, n (%) | Chi-square or Fisher's exact test as appropriate | 3 (6) | 2 (15) | 0.28 | 3 (6) | 3 (21) | 0.12 | 3 (6) | 3 (20) | 0.15 | 3 (7) | 1 (8) | 1.00 |
| | Necrotizing enterocolitis, grade 2-3, n (%) | Chi-square or Fisher's exact test as appropriate | 2 (4) | 1 (8) | 0.51 | 2 (4) | 2 (14) | 0.21 | 1 (2) | 3 (20) | **0.04** | 3 (7) | 2 (15) | 0.31 |
| | Days on antibiotics, mean (SD) | t-test for independent samples | 24 (14) | 35 (17) | **0.01** | 24 (13) | 34 (17) | **0.02** | 24 (13) | 35 (16) | **0.01** | 25 (14) | 34 (15) | **0.04** |

| | | | week 1 | | | week 2 | | | week 3 | | | week 4 | | |
|---|---|---|---|---|---|---|---|---|---|---|---|---|---|---|
| | **Variables** | **Statistical test** | **Normal (n=48)** | **Impairment (n=42)** | **p value** | **Normal (n=50)** | **Impairment (n=42)** | **p value** | **Normal (n=43)** | **Impairment (n=45)** | **p value** | **Normal (n=44)** | **Impairment (n=41)** | **p value** |
| **NDI** | Gestational age, weeks, mean (SD) | t-test for independent samples | 25,8 (1,0) | 25,0 (1,3) | **<0,001** | 25,9 (1,0) | 25,0 (1,3) | **<0.001** | 26,0 (0,9) | 25,0 (1,3) | **<0,001** | 25,9 (1,0) | 25,0 (1,2) | **<0,001** |
| | Birth weight, g, mean (SD) | t-test for independent samples | 796 (126) | 682 (120) | **<0,001** | 799 (128) | 686 (121) | **<0.001** | 807 (114) | 688 (121) | **<0,001** | 799 (125) | 695 (117) | **<0,001** |
| | Birth weight z-score, mean (SD) | t-test for independent samples | -0.9 (1,1) | -1,1 (1,3) | 0.21 | -0,9 (1,1) | -1,1 (1,2) | 0.27 | -0,9 (1,0) | -1,1 (1,3) | 0.45 | -1,0 (1,1) | -1,0 (1,1) | 0.86 |
| | Birth length, cm, mean (SD) | t-test for independent samples | 33,6 (2,5) | 31,6 (2,1) | **<0,001** | 33,6 (2,5) | 31,6 (2,1) | **<0,001** | 33,9 (2,2) | 31,7 (2,1) | **<0,001** | 33,6 (2,5) | 31,9 (2,0) | **<0,001** |
| | Birth length, z-score, mean (SD) | t-test for independent samples | -1,1 (1,8) | -1,6 (1,6) | 0.07 | -1,1 (1,8) | -1,6 (1,6) | 0.09 | -1,0 (1,5) | -1,6 (1,6) | 0.06 | -1,1 (1,8) | -1,4 (1,4) | 0.44 |
| | Birth head circumference, cm, mean (SD) | t-test for independent samples | 23,3 (1,3) | 22,4 (1,4) | **0.001** | 23,4 (1,4) | 22,5 (1,5) | **<0,001** | 25,6 (1,3) | 22,5 (1,5) | **<0,001** | 23,5 (1,4) | 22,5 (1,4) | **0.001** |
| | Birth head circumference z-score, mean (SD) | t-test for independent samples | -0,8 (0,7) | -0,7 (0,7) | 0.28 | -0,8 (0,8) | -0,7 (0,7) | 0.27 | -0,8 (0,7) | -0,7 (0,7) | 0.55 | -0,8 (0,8) | -0,7 (0,7) | 0.66 |
| | Apgar at 10 minutes, mean (SD) | t-test for independent samples | 8,2 (1,7) | 7,6 (1,8) | 0.08 | 8,1 (1,8) | 7,6 (1,7) | 0.10 | 8,4 (1,4) | 7,6 (1,7) | **0.02** | 8,1 (1,8) | 7,3 (1,8) | **0.03** |
| | SGA, n (%) | Chi-square or Fisher's exact test as appropriate | 12 (25) | 8 (19) | 0.50 | 13 (26) | 7 (17) | 0.28 | 11 (26) | 8 (18) | 0.44 | 13 (30) | 6 (15) | 0.10 |
| | Female sex, n (%) | Chi-square or Fisher's exact test as appropriate | 25 (52) | 17 (40) | 0.27 | 26 (52) | 17 (40) | 0.27 | 24 (56) | 18 (40) | 0.14 | 23 (52) | 14 (34) | 0.09 |
| | Infants from multiple pregnancy, n (%) | Chi-square or Fisher's exact test as appropriate | 18 (38) | 12 (29) | 0.37 | 21 (42) | 15 (36) | 0.67 | 19 (44) | 16 (36) | 0.41 | 20 (46) | 13 (32) | 0.19 |
| | Prenatal steroids administered, n (%) | Chi-square or Fisher's exact test as appropriate | 46 (96) | 42 (100) | 0.50 | 48 (96) | 42 (100) | 0.50 | 42 (98) | 45 (100) | 0.49 | 43 (98) | 41 (100) | 1.00 |
| | Chorioamnionitis, n (%) | Chi-square or Fisher's exact test as appropriate | 14 (29) | 6 (14) | 0.09 | 15 (30) | 8 (19) | 0.23 | 13 (30) | 9 (20) | 0.27 | 13 (30) | 7 (17) | 0.18 |
| | Cesarean section, n (%) | Chi-square or Fisher's exact test as appropriate | 33 (69) | 25 (60) | 0.32 | 35 (70) | 25 (60) | 0.29 | 33 (77) | 29 (64) | 0.21 | 33 (75) | 26 (63) | 0.25 |
| | Smoking in family at inclusion, n (%) | Chi-square or Fisher's exact test as appropriate | 1 (2) | 6 (14) | 0.05 | 1 (2) | 5 (12) | 0.09 | 0 (0) | 5 (11) | 0.06 | 1 (2) | 6 (15) | 0.05 |
| | Inclusion site | Chi-square or Fisher's exact test as appropriate | | | | | | | | | | | | |
| | Linkoping, n (%) | | 10 (21) | 22 (54) | **0.002** | 11 (22) | 22 (52) | **0.002** | 10 (23) | 22 (49) | **0.01** | 9 (20) | 21 (51) | **0.004** |
| | Stockholm, n (%) | | 38 (79) | 20 (48) | | 39 (78) | 28 (48) | | 33 (77) | 23 (51) | | 35 (80) | 20 (49) | |
| | **Neonatal hospital care** | | | | | | | | | | | | | |
| | Bronchopulmonary dysplasia, n (%) | Chi-square or Fisher's exact test as appropriate | 20 (42) | 32 (76) | **<0,001** | 21 (42) | 32 (76) | **<0,001** | 17 (40) | 35 (78) | **<0,001** | 18 (50) | 34 (83) | **<0,001** |
| | Sepsis, n (%) | Chi-square or Fisher's exact test as appropriate | 11 (23) | 14 (33) | 0.27 | 12 (24) | 13 (31) | 0.46 | 10 (23) | 16 (36) | 0.21 | 11 (25) | 9 (22) | 0.74 |
| | Retinopathy of prematurity, grade 3-5, n (%) | Chi-square or Fisher's exact test as appropriate | 3 (6) | 12 (29) | **0.01** | 3 (6) | 9 (21) | 0.03 | 3 (7) | 10 (22) | **0.04** | 3 (7) | 11 (27) | 0.01 |
| | Intraventricular haemorrhage, grade 3-4, n (%) | Chi-square or Fisher's exact test as appropriate | 3 (6) | 6 (14) | 0.30 | 3 (6) | 8 (19) | 0.06 | 3 (7) | 8 (18) | 0.13 | 3 (7) | 5 (12) | 0.47 |
| | Necrotizing enterocolitis, grade 2-3, n (%) | Chi-square or Fisher's exact test as appropriate | 1 (2) | 3 (7) | 0.34 | 1 (2) | 4 (10) | 0.17 | 0 (0) | 5 (11) | 0.06 | 2 (4) | 4 (10) | 0.42 |
| | Days on antibiotics, mean (SD) | t-test for independent samples | 20 (10) | 33 (15) | **<0,001** | 20 (10) | 34 (15) | **<0,001** | 20 (10) | 34 (15) | **<0,001** | 20 (11) | 33 (14) | **<0,001** |

**Supplementary Table S3.** Probiotic supplementation in relation to Bayley-II scores (language (T-test), cognition (Mann-Whitney U-test), and motor (T-test)), Alpha-diversity (Shannon, richness, and evenness), Beta-diversity (ANOSIM test), and differential abudnacne (ANCOM test).
SD: standard deviation; lfc: log fold change placebo vs *L. reuteri.*

| Supplementation | Timepoint | N | Bayley-III score | | | | | | Chi-squre | | |
|---|---|---|---|---|---|---|---|---|---|---|---|
| | | | Outcome | Mean | Median | SD | p-value | q-value | Impairment | Normal | p-value |
| Placebo | w1 | 36 | Language | 82 | 86 | 14.1 | 0.05 | 0.06 | 17 | 19 | 1 |
| *L. reuteri* | | 31 | | 89 | 86 | 15.6 | | | 14 | 17 | |
| Placebo | w2 | 36 | | 82 | 86 | 14.1 | 0.05 | 0.06 | 16 | 20 | 1 |
| *L. reuteri* | | 31 | | 90 | 86 | 15.9 | | | 13 | 18 | |
| Placebo | w3 | 35 | | 81 | 86 | 14.4 | 0.05 | 0.06 | 17 | 18 | 0.9761 |
| *L. reuteri* | | 31 | | 88 | 86 | 16.0 | | | 14 | 17 | |
| Placebo | w4 | 30 | | 82 | 86 | 14.5 | 0.09 | 0.09 | 14 | 16 | 0.9091 |
| *L. reuteri* | | 31 | | 89 | 86 | 16.1 | | | 13 | 18 | |
| Placebo | w1 | 40 | Cognition | 89 | 95 | 15.5 | 0.92 | NA | 12 | 28 | 1 |
| *L. reuteri* | | 31 | | 90 | 90 | 13.4 | | | 10 | 21 | |
| Placebo | w2 | 39 | | 89 | 95 | 15.9 | 0.87 | NA | 12 | 27 | 1 |
| *L. reuteri* | | 31 | | 90 | 90 | 14.3 | | | 10 | 21 | |
| Placebo | w3 | 38 | | 87 | 95 | 16.0 | 0.97 | NA | 13 | 25 | 1 |
| *L. reuteri* | | 31 | | 89 | 90 | 14.0 | | | 10 | 21 | |
| Placebo | w4 | 33 | | 89 | 95 | 15.6 | 0.84 | NA | 10 | 23 | 1 |
| *L. reuteri* | | 32 | | 91 | 90 | 13.8 | | | 9 | 23 | |
| Placebo | w1 | 37 | Motor | 87 | 88 | 15.1 | 0.22 | NA | 8 | 29 | 1 |
| *L. reuteri* | | 25 | | 92 | 88 | 17.1 | | | 5 | 20 | |
| Placebo | w2 | 37 | | 87 | 88 | 15.0 | 0.28 | NA | 8 | 29 | 1 |
| *L. reuteri* | | 26 | | 92 | 88 | 17.1 | | | 6 | 20 | |
| Placebo | w3 | 36 | | 86 | 88 | 16.2 | 0.13 | NA | 9 | 27 | 1 |
| *L. reuteri* | | 25 | | 92 | 88 | 17.7 | | | 6 | 19 | |
| Placebo | w4 | 31 | | 86 | 88 | 16.0 | 0.31 | NA | 7 | 24 | 1 |
| *L. reuteri* | | 27 | | 90 | 88 | 16.9 | | | 6 | 21 | |

| Supplementation | Timepoint | Alpha-diverstiy | | | | | | Beta-diversity | | ANCOM | | NDI | | |
|---|---|---|---|---|---|---|---|---|---|---|---|---|---|---|
| | | Metric | Mean | Median | SD | p-value | q-value | Anosim significance | Anosim StatisticR | taxon | lfc | Impairment | Normal | Chi2 p-value |
| Placebo | w1 | Shannon | 1.0 | 1.0 | 0.4 | 3.4E-12 | 1.3E-11 | 0.00 | 0.18 | Lactobacillus | 6.841508 | 22 | 25 | 1 |
| *L. reuteri* | | | 1.7 | 1.6 | 0.4 | | | | | | | 20 | 23 | |
| Placebo | w2 | | 1.1 | 1.1 | 0.5 | 1.0E-07 | 2.1E-07 | 0.00 | 0.30 | Lactobacillus | 6.289069 | 22 | 26 | 1 |
| *L. reuteri* | | | 1.7 | 1.7 | 0.4 | | | | | | | 20 | 24 | |
| Placebo | w3 | | 1.2 | 1.2 | 0.6 | 1.2E-02 | 1.6E-02 | 0.00 | 0.15 | Lactobacillus | 3.921682 | 23 | 22 | 1 |
| *L. reuteri* | | | 1.5 | 1.5 | 0.5 | | | | | | | 22 | 21 | |
| Placebo | w4 | | 1.3 | 1.4 | 0.5 | 3.7E-02 | 3.7E-02 | 0.44 | 0.00 | Lactobacillus | 4.117803 | 21 | 20 | 0.753 |
| *L. reuteri* | | | 1.6 | 1.6 | 0.5 | | | | | | | 20 | 24 | |
| Placebo | w1 | Richness | 12 | 10 | 6.2 | 3.3E-05 | 1.3E-04 | | | | | | | |
| *L. reuteri* | | | 18 | 17 | 5.6 | | | | | | | | | |
| Placebo | w2 | | 20 | 20 | 7.4 | 2.7E-03 | 4.5E-03 | | | | | | | |
| *L. reuteri* | | | 25 | 25 | 7.8 | | | | | | | | | |
| Placebo | w3 | | 20 | 18 | 7.3 | 3.4E-03 | 4.5E-03 | | | | | | | |
| *L. reuteri* | | | 25 | 24 | 7.2 | | | | | | | | | |
| Placebo | w4 | | 23 | 23 | 7.7 | 3.7E-01 | 3.7E-01 | | | | | | | |
| *L. reuteri* | | | 24 | 24 | 7.6 | | | | | | | | | |
| Placebo | w1 | Evenness | 0.4 | 0.4 | 0.2 | 2.7E-05 | 5.5E-05 | | | | | | | |
| *L. reuteri* | | | 0.6 | 0.6 | 0.1 | | | | | | | | | |
| Placebo | w2 | | 0.4 | 0.4 | 0.2 | 2.8E-06 | 1.1E-05 | | | | | | | |
| *L. reuteri* | | | 0.5 | 0.5 | 0.1 | | | | | | | | | |
| Placebo | w3 | | 0.4 | 0.4 | 0.2 | 3.3E-02 | 3.3E-02 | | | | | | | |
| *L. reuteri* | | | 0.5 | 0.5 | 0.2 | | | | | | | | | |
| Placebo | w4 | | 0.4 | 0.5 | 0.2 | 3.0E-02 | 3.3E-02 | | | | | | | |
| *L. reuteri* | | | 0.5 | 0.5 | 0.2 | | | | | | | | | |

**Supplementary Table S4**. Estimated microbial mediation effect (SparseMCMM).

| | | Estimated P-value | | Estimated causal effects | | |
|---|---|---|---|---|---|---|
| **Outcome** | **Time point** | **OME** | **CME** | **ME** | **DE** | **TE** |
| Language | w1 | 0.396 | 0.416 | -66.018 | 71.578 | 5.561 |
| | w2 | 0.446 | 0.089 | 14.917 | -2.670 | 12.247 |
| | w3 | 0.386 | 0.446 | -62.164 | 146.885 | 84.721 |
| | w4 | 0.366 | 0.347 | 52.115 | -53.591 | -1.475 |
| Cognition | w1 | 0.396 | 0.416 | 32.054 | -32.441 | -0.387 |
| | w2 | 0.475 | 0.317 | -12.057 | 11.611 | -0.446 |
| | w3 | 0.455 | 0.485 | 30.182 | -16.671 | 13.511 |
| | w4 | 0.426 | 0.406 | -42.678 | 41.848 | -0.830 |
| Motor | w1 | 0.356 | 0.376 | 34.225 | -35.543 | -1.318 |
| | w2 | 0.545 | 0.465 | -8.807 | 19.385 | 10.579 |
| | w3 | 0.554 | 0.545 | 35.447 | -21.397 | 14.050 |
| | w4 | 0.782 | 0.604 | 3.773 | 15.194 | 18.967 |

OME: tests the overall mediation effect of the microbiome community
CME: test whether at least one taxon has a mediation effect.
ME: estimates of the overall microbial mediation effect
DE: estimates of the direct treatment effect.
TE: estimates of the total treatment effect.

**Supplementary Table S5.** Causal Mediation Analysis with alpha-diversity as the mediator.

| | | week 1 | | | | week 2 | | | | week 3 | | | | week 4 | | | |
|---|---|---|---|---|---|---|---|---|---|---|---|---|---|---|---|---|---|
| | | **Estimate** | **95% CI Lower** | **95% CI Upper** | **p-value** | **Estimate** | **95% CI Lower** | **95% CI Upper** | **p-value** | **Estimate** | **95% CI Lower** | **95% CI Upper** | **p-value** | **Estimate** | **95% CI Lower** | **95% CI Upper** | **p-value** |
| **Diversity** | Total Effect | 7.13 | 0.23 | 14.94 | 0.05 | 7.26 | 0.21 | 15.46 | 0.04 | 8.08 | 0.66 | 15.32 | 0.03 | 6.75 | -0.94 | 14.67 | 0.09 |
| | ACME | 1.42 | -4.79 | 10.78 | 0.64 | 1.63 | -2.62 | 8.68 | 0.42 | 0.82 | -2.65 | 4.39 | 0.75 | 0.34 | -2.31 | 3.86 | 0.87 |
| | ADE | 5.70 | -4.19 | 14.44 | 0.23 | 5.64 | -2.51 | 13.97 | 0.18 | 7.26 | -0.66 | 15.67 | 0.07 | 6.41 | -1.14 | 14.33 | 0.10 |
| | Prop. Mediated | 0.20 | -1.52 | 2.33 | 0.64 | 0.22 | -0.71 | 1.77 | 0.45 | 0.10 | -0.56 | 0.99 | 0.76 | 0.05 | -0.91 | 0.92 | 0.86 |
| **Richness** | Total Effect | 7.23 | 0.26 | 14.47 | 0.04 | 7.26 | 0.21 | 15.46 | 0.04 | 7.35 | 0.61 | 15.25 | 0.03 | 6.34 | -0.61 | 14.36 | 0.07 |
| | ACME | 1.78 | -0.72 | 5.67 | 0.18 | 1.63 | -2.62 | 8.68 | 0.42 | 1.32 | -0.60 | 4.53 | 0.15 | 0.38 | -1.45 | 3.15 | 0.51 |
| | ADE | 5.46 | -1.45 | 12.71 | 0.13 | 5.64 | -2.51 | 13.97 | 0.18 | 6.03 | -1.68 | 14.60 | 0.13 | 5.96 | -1.31 | 14.01 | 0.12 |
| | Prop. Mediated | 0.25 | -0.24 | 1.32 | 0.20 | 0.22 | -0.71 | 1.77 | 0.45 | 0.18 | -0.12 | 1.63 | 0.18 | 0.06 | -0.46 | 1.07 | 0.54 |
| **Evenness** | Total Effect | 7.08 | -0.15 | 16.61 | 0.06 | 6.92 | -0.22 | 16.65 | 0.06 | 7.63 | 0.32 | 15.70 | 0.04 | 6.68 | -1.65 | 15.60 | 0.11 |
| | ACME | 0.75 | -4.17 | 9.95 | 0.68 | 0.36 | -3.44 | 9.30 | 0.71 | -0.03 | -3.43 | 4.68 | 0.96 | 0.11 | -3.53 | 4.85 | 0.97 |
| | ADE | 6.33 | -2.35 | 14.63 | 0.14 | 6.57 | -1.18 | 14.70 | 0.09 | 7.66 | -0.09 | 15.91 | 0.05 | 6.57 | -0.96 | 14.49 | 0.09 |
| | Prop. Mediated | 0.11 | -1.42 | 1.69 | 0.68 | 0.05 | -0.91 | 1.18 | 0.70 | 0.00 | -0.83 | 0.79 | 0.96 | 0.02 | -1.48 | 1.13 | 0.92 |

Total Effect: indicates whether probiotic supplementation affects Language development.
ACME: average Causal Mediation Effect (indirect effect), indicates whether alpha-diversity had a mediating effect.
ADE: Average Direct Effect.

**Supplementary Table S6. Loading weights of the ASVs on PLS-DA component 1.**
The table shows the ASV signature that characterize each group (normal or impairment). See table in excel format.

**Supplementry Table S7**. Classification accuracy measures. Apparent AUC (AUC of the signature applied to the same data that was used to generate the model), mean cross-validation (cv) AUC, standard deviation (sd) cv-AUC.

| | **apparent AUC** | **mean cv-AUC** | **sd cv-AUC** |
|---|---|---|---|
| Language | 0.65 | 0.55 | 0.04 |
| Cognition | 0.63 | 0.49 | 0.10 |
| Motor | 0.80 | 0.74 | 0.02 |
| NDI | 0.63 | 0.58 | 0.05 |

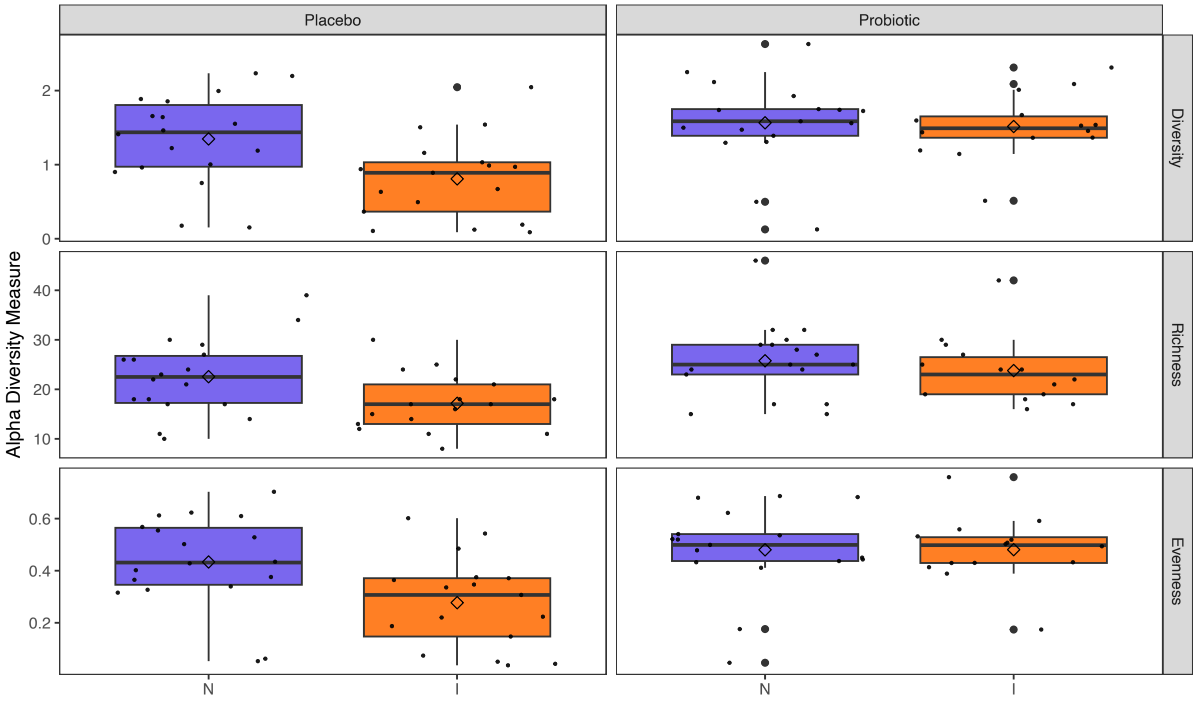

**Supplementary Figure S1.** Gut microbiota alpha-diversity of ELBW-EPT infants with normal (N) or impaired (I) language development at 2 years of age.

Boxplots (median with 25% and 75% percentiles and 1.5x the interquartile range; diamond shape depicts the mean) showing the a-diversity (Shannon index), richness (observed ASVs), and evenness (Pielou’s evenness index) from 1 week to 4 weeks of life.

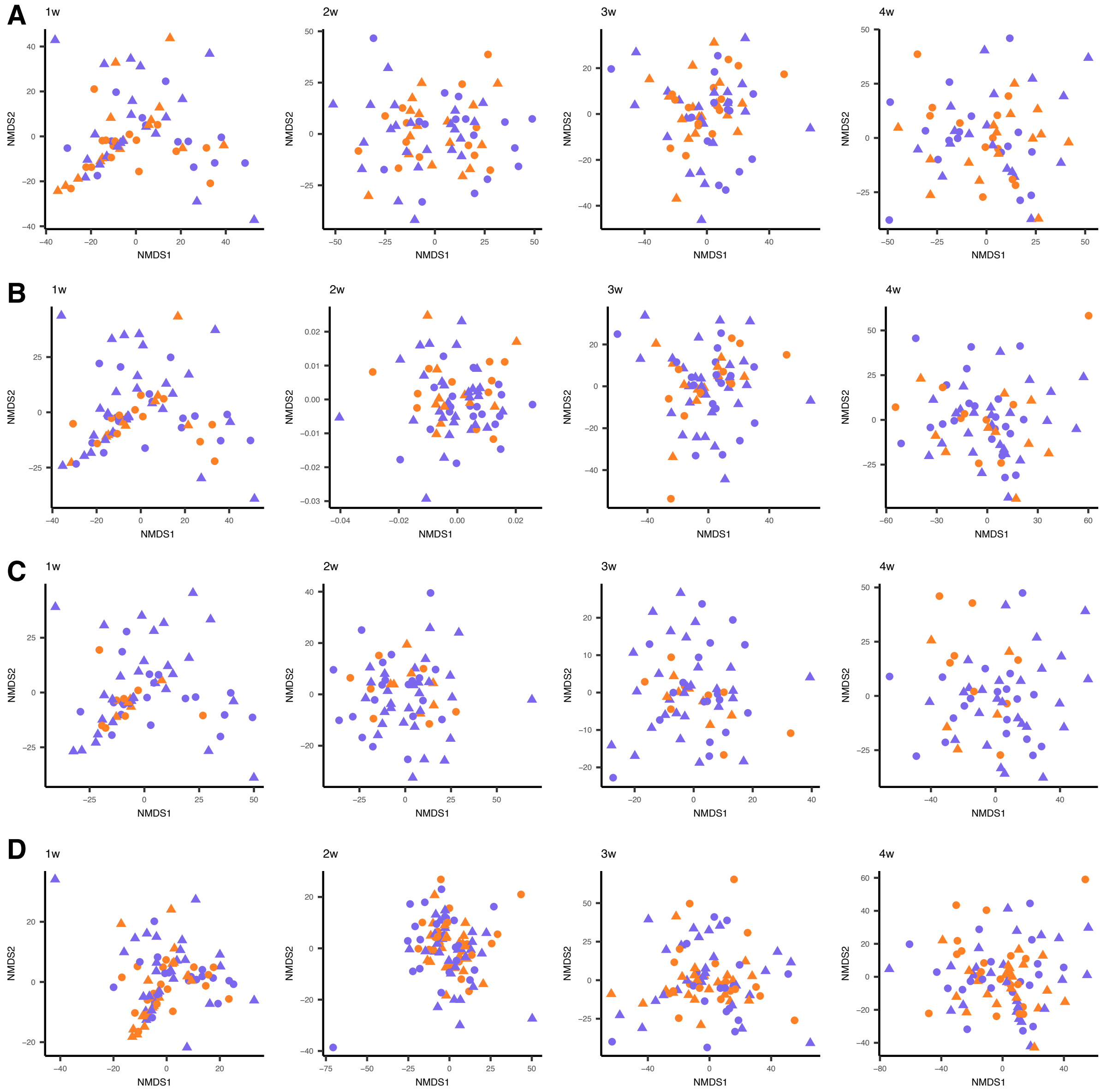

**Supplementary Figure S2**. Non-metric multidimensional scaling (NMDS) of the bacterial community composition from 1week (w) to 4 weeks (w) of life across ELBW-EPT infants with normal vs impaired language (A), cognition (B), and motor (C) development, overall neurodevelopment (D) at 2 years of age. Purple: normal development, orange: impaired development.

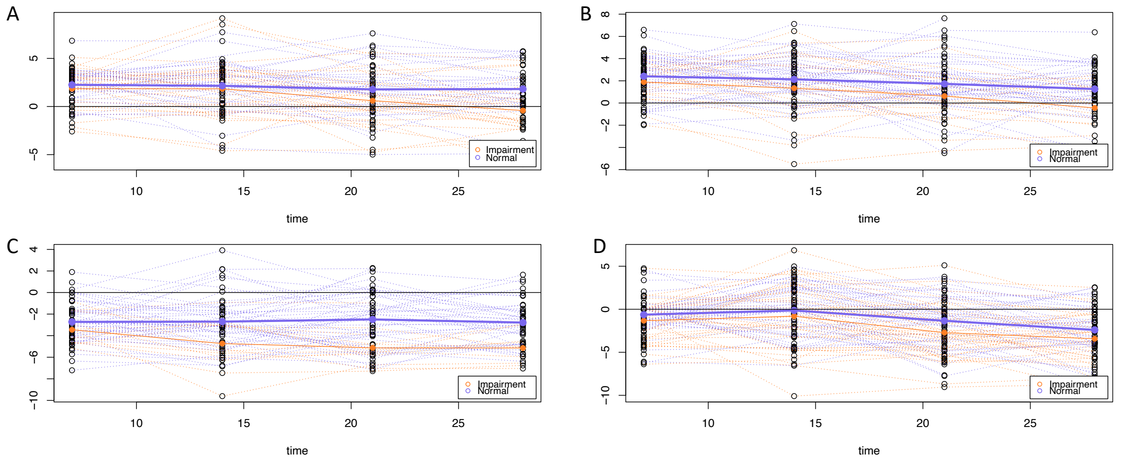

**Supplementary Figure S3.** Microbial signature trajectories. Mean relative abundance of the microbial signature discriminative for normal and impaired groups for language development (A), cognition development (B), motor development (C), and overall neurodevelopment impairment (D).
